# Spectrum of somatic mutational features of colorectal tumors in ancestrally diverse populations

**DOI:** 10.1101/2024.03.11.24303880

**Authors:** Marco Matejcic, Jamie K. Teer, Hannah J. Hoehn, Diana B. Diaz, Kritika Shankar, Jun Gong, Nathalie T. Nguyen, Nicole Lorona, Domenico Coppola, Clifton Fulmer, Ozlen Saglam, Kun Jiang, Douglas Cress, Teresita Muñoz-Antonia, Idhaliz Flores, Edna Gordian, José A. Oliveras Torres, Seth I. Felder, Julian A. Sanchez, Jason Fleming, Erin M. Siegel, Jennifer A. Freedman, Julie Dutil, Mariana C. Stern, Brooke L. Fridley, Jane C. Figueiredo, Stephanie L. Schmit

**Affiliations:** Department of Biostatistics and Bioinformatics, H. Lee Moffitt Cancer Center & Research Institute, Tampa, FL; Department of Cancer Epidemiology, H. Lee Moffitt Cancer Center & Research Institute, Tampa, FL; Non-Therapeutic Research Office, H. Lee Moffitt Cancer Center & Research Institute, Tampa, FL; Genomic Medicine Institute, Lerner Research Institute, Cleveland Clinic, Cleveland, OH; Department of Medicine, Samuel Oschin Comprehensive Cancer Institute, Cedars-Sinai Medical Center, Los Angeles, CA; Department of Anatomic Pathology, H. Lee Moffitt Cancer Center & Research Institute, Tampa, FL; Department of Pathology, Robert J. Tomsich Pathology and Laboratory Medicine Institute, Cleveland Clinic Foundation, Cleveland, OH; Department of Molecular Oncology, H. Lee Moffitt Cancer Center & Research Institute, Tampa, FL; Puerto Rico Biobank, H. Lee Moffitt Cancer Center & Research Institute, Tampa, Florida and Ponce Health Sciences University, Ponce, PR; Department of Gastrointestinal Oncology, H. Lee Moffitt Cancer Center & Research Institute, Tampa, FL; Department of Medicine, Division of Medical Oncology, Duke University School of Medicine, Durham, NC; Duke Cancer Institute, Durham, NC; Cancer Biology Division, Ponce Research Institute, Ponce Health Sciences University, Ponce, Puerto Rico; Department of Preventive Medicine, Keck School of Medicine, University of Southern California/Norris Comprehensive Cancer Center, Los Angeles, CA, USA; Population and Cancer Prevention Program, Case Comprehensive Cancer Center, Cleveland, OH

**Keywords:** Ancestry, Colorectal Cancer, Genomics, Hispanic, Latin*

## Abstract

Ancestrally diverse and admixed populations, including the Hispanic/Latino/a/x/e community, are underrepresented in cancer genetic and genomic studies. Leveraging the Latino Colorectal Cancer Consortium, we analyzed whole exome sequencing data on tumor/normal pairs from 718 individuals with colorectal cancer (128 Latino, 469 non-Latino) to map somatic mutational features by ethnicity and genetic ancestry.

Global proportions of African, East Asian, European, and Native American ancestries were estimated using ADMIXTURE. Associations between global genetic ancestry and somatic mutational features across genes were examined using logistic regression.

*TP53*, *APC*, and *KRAS* were the most recurrently mutated genes. Compared to non-Latino individuals, tumors from Latino individuals had fewer *KRAS* (OR=0.64, 95%CI=0.41-0.97, p=0.037) and *PIK3CA* mutations (OR=0.55, 95%CI=0.31-0.98, p=0.043). Genetic ancestry was associated with presence of somatic mutations in 39 genes (FDR-adjusted LRT p<0.05). Among these genes, a 10% increase in African ancestry was associated with significantly higher odds of mutation in *KNCN* (OR=1.34, 95%CI=1.09–1.66, p=5.74×10^-3^) and *TMEM184B* (OR=1.53, 95%CI=1.10–2.12, p=0.011). Among RMGs, we found evidence of association between genetic ancestry and mutation status in *CDC27* (LRT p=0.0084) and between *SMAD2* mutation status and AFR ancestry (OR=1.14, 95%CI=1.00-1.30, p=0.046). Ancestry was not associated with tumor mutational burden. Individuals with above-average Native American ancestry had a lower frequency of microsatellite instable (MSI-H) vs microsatellite stable tumors (OR=0.45, 95%CI=0.21-0.99, p=0.048).

Our findings provide new knowledge about the relationship between ancestral haplotypes and somatic mutational profiles that may be useful in developing precision medicine approaches and provide additional insight into genomic contributions to cancer disparities.

**Significance:** Our data in ancestrally diverse populations adds essential information to characterize mutational features in the colorectal cancer genome. These results will help enhance equity in the development of precision medicine strategies.

## Introduction

Therapeutic options for cancer are increasingly being selected based on the presence of specific somatic mutations and other genetic and molecular features of tumors. There are numerous examples of somatic features that can be targeted with existing FDA-approved therapies for precision oncology (1). For colorectal cancer (CRC), molecularly-driven therapies are rapidly expanding and currently include immune checkpoint inhibitors for microsatellite instable (MSI-H) tumors, small molecule inhibitors (adagrasib and sotorasib) for mutant *KRAS* G12C tumors, anti-epidermal growth factor receptor (EGFR) inhibitors (cetuximab and panitumumab) for *RAS*/*BRAF* wildtype left-sided primary tumors, BRAF inhibitors with anti-EGFR inhibitors for *BRAF* V600E mutant tumors, NTRK inhibitors for tumors with *NTRK* fusions, and HER2-directed therapies for HER2-amplified tumors (2). Novel molecularly-driven therapies for CRC in the pipeline include combination immunotherapies to overcome the immune “cold” nature of microsatellite stable (MSS) tumors, small molecule inhibitors against *KRAS* G12D and other *KRAS* mutations, and therapies targeting the MAPK pathway (2). As precision oncology approaches are poised to revolutionize cancer care, even greater attention is needed to ensure that their benefits are equitably applicable to all patients.

For decades, disparities in treatment response and cancer-related mortality have been reported across populations by factors such as socioeconomics, geography and racial and ethnic identities (3–5). Structural inequities and social injustices clearly impact prognosis and contribute to these persistent disparities (6,7). Interrelated with self-identified race and ethnicity is genetic ancestry, which measures inherited genetic variation linked to human migration patterns (8). Early evidence suggests that germline genetic variation may influence the spectrum of somatic alterations that accumulate during tumorigenesis (9,10). Emerging data report differences by ancestry in the frequency of somatic alterations and molecular features. For example, Yuan et al. leveraged data from The Cancer Genome Atlas (TCGA) to show that selected tumors (i.e., breast, head and neck and endometrium) occurring in individuals with African ancestry have higher levels of chromosomal instability and fewer genetic alterations in the PI3K pathway when compared to individuals of European ancestry (11).

Admixed populations, such as Hispanic/Latino/a/x/e (henceforth referred to as Latino) individuals, inherited alleles from multiple ancestral groups (i.e., primarily Native American, European, and African) (12), and have potential to offer unique insight into how ancestry shapes the somatic mutational landscape of tumors. These data have lagged behind those generated for other populations, in part due to limitations of existing resources including TCGA, which includes fewer than 3% of tumors from Latino individuals and <0.5% from Indigenous American individuals (13). However, novel efforts to build the infrastructure for genomic research in Hispanic/Latin American populations have begun to address this gap and create complementary resources and generate important findings. In 2021, Carrot-Zhang et al. (14) reported results on the contributions of genetic ancestry in lung tumors among admixed Latin American populations. Striking associations between Native American (NAT) ancestry and somatic tumor features, including tumor mutational burden (TMB) and specific driver mutations in *EGFR*, *KRAS* and *STK11*, were reported. In 2023, Ding et al. (15) reported results from a Californian study of Hispanic women with breast cancer, showing a similar prevalence of mutations in established driver genes including *PIK3CA*, *TP53*, *GATA3*, *MAP3K1*, *CDH1*, *CBFB*, *PTEN* and *RUNX1* and recurrent amplifications in breast cancer drivers including *MYC*, *FGFR1*, *CCND1* and *ERBB2* as compared to samples from non-Hispanic White participants in TCGA. Importantly, they identified a COSMIC signature in a significant fraction of tumors, which was observed (but not reported) in the Romero-Cordoba et al. (16) study among Hispanic women with breast cancer living in Mexico. The latter study also identified a high frequency (8%) of E17K-activating *AKT1* mutations in Mexican women, who may benefit from AKT inhibitors (16). These studies recapitulate known alterations observed in other populations, but importantly, also reveal novel associations. Achieving representation of tumors from patients of diverse populations will allow greater opportunity to characterize variation of the tumor genome and aid precision medicine approaches equitably.

Here, we address the underrepresentation of Latino individuals in CRC genomic research. We characterize the whole-exome mutational landscapes of tumors from 718 CRC patients, including 128 Latino patients from the Latino Colorectal Cancer Consortium (LC3) and 469 non-Latino patients. We examine associations between gene-level mutational status and both global genetic ancestry and race/ethnicity, two complementary aspects of identity. We also examine the frequency of clinically relevant features: defective mismatch repair (dMMR)/MSI-H status (henceforth referred to as MSI status) and TMB.

## Methods

### Study populations and biospecimen collection

The Latino Colorectal Cancer Consortium (LC3) is a partnership between several epidemiologic studies and biobanks to promote harmonized infrastructure for the study of CRC disparities among heterogeneous Latino communities and between the Latino population and other racial and ethnic groups. It includes patient data and biospecimens from three underlying studies: the Hispanic Colorectal Cancer Study (HCCS) (17), the Puerto Rico Biobank (PRBB) (18), and Total Cancer Care (TCC) at H. Lee Moffitt Cancer Center and Research Institute (19) and from TCGA (https://portal.gdc.cancer.gov). Detailed information on recruitment and selection of study participants is provided in the **Supplementary Materials**. All participants either provided written informed consent or a HIPAA waiver of consent was obtained from the appropriate Institutional Review Board. The study protocol was approved by the Institutional Review Boards at Cleveland Clinic (LC3 central coordinating center), Cedars Sinai Medical Center (HCCS), H. Lee Moffitt Cancer Center and Research Institute (TCC; PRBB), and Ponce Research Institute (PRBB).

### Patient characteristics and clinical data

Socio-demographic data included sex, ethnicity (Latino, non-Latino), and race (American Indian, Asian, Black, White, Other). Here, we use the terms race and ethnicity to describe non-biological social categories based on cultural traditions, nativity, language, and religion. Of the 597 patients with available ethnicity information, 128 were Latino (123 from LC3; 5 from TCGA) and 469 non-Latino (173 from TCC; 296 from TCGA). Latino individuals were included in the Latino ethnic group regardless of race. Additional demographic and clinical data were abstracted from medical records or cancer registries and included age at diagnosis, tumor location, stage at diagnosis, and MSI status. The distribution of patient and tumor characteristics for the study participants stratified by ethnicity are presented in **Supplementary Table 1**.

### Sequencing analysis and quality control

Tumor DNA was derived either from fresh frozen or FFPE (formalin-fixed paraffin-embedded) tissue; germline/normal DNA was derived from blood, saliva, or normal colorectal tissue. Whole exome sequencing was performed for tumors and matched normal samples using protocols for DNA extraction, exome capture, and sequencing specific to each contributing study (**Supplementary Table 2**). Sequence reads were aligned to the reference human genome (hs37d5) with Burrows-Wheeler Aligner (BWA) (20) and refined with the Genome Analysis ToolKit (GATK) (21) and Picard (http://picard.sourceforge.net/).

Tumor-specific somatic mutations from matched pairs were identified using a bioinformatic pipeline including Strelka (22) and MuTect (23). Mutations observed as PASS in Strelka or observed at all in Strelka and PASS in MuTect were retained for analysis. To further remove false positive mutations, any mutation detected from our cohort that exceeded a 1% allele frequency in all samples based on the 1KGP Phase 3 dataset was removed. Inherited variants were called using GATK UnifiedGenotyper. All germline variants and mutations were filtered on the intersection of all target regions (padded to include intron/exon junctions). Median depth of sequence read coverage was 116x in tumor and 108x in germline samples. Proper tumor/normal pairing was confirmed by comparing inherited variant calls at common single nucleotide polymorphism (SNP) positions using TimeAttackGenComp (24) (https://github.com/teerjk/TimeAttackGenComp). Mutation allele frequency was compared between tumor samples and normal samples to filter out artifact mutations: mutations were considered passing with allele frequency skew in tumor vs normal of ≥2 at positions with depth of coverage ≥10 in at least 80% of tumor and normal samples (see details in the **Supplementary Materials**).

### Mutation annotation

Mutations were annotated with gene context, predicted functional effect, and population allele frequencies from the 1KGP (25) (http://www.internationalgenome.org/) using ANNOVAR May-2012 (26). Only the mutations resulting in a change in protein sequence and located in gene bodies based on the GRCh37 assembly were included in the association analyses (i.e., synonymous, intronic, 5’ UTR and 3’ UTR mutations were excluded). In lollipop-style mutation diagrams, only mutations with protein change predictions are presented.

### Global ancestry estimation

The 1KGP and the PAGE Study (http://www.pagestudy.org/) were used as reference datasets for genetic ancestry estimation (25,27). Here, we use the term genetic ancestry to describe the proportion of an individual’s genome derived from major continental ancestral populations based on sequence similarity; specifically, we utilize EAS, AFR, NAT and EUR to denote Asian, African, Native American, and European continental ancestral populations, respectively. The distribution of reference samples by major population group in the 1KGP and the PAGE Study is presented in **Supplementary Table 3**. To remove admixed samples and improve accuracy in global ancestry estimation, reference samples from underrepresented populations as well as samples without a major ancestral component (≤98% of the total) were excluded. After removing highly admixed individuals, genetic markers from the 1KGP (n=1,668) and the PAGE Study (n=1,047) were merged with germline variants from the study participants and were used as input data for global ancestry estimation. We performed a supervised analysis in ADMIXTURE-1.3.0 to estimate the ancestral components in each individual (28). Non-admixed reference samples from 1KGP and the PAGE Study were labeled with one of the four major continental ancestral populations to better estimate ancestry in our study samples (LC3 and TCGA). The model was run using K=4, which is the minimum value that represented the 4 major continental ancestral components in US Latino individuals (EAS, AFR, NAT, EUR). Details on data processing are provided in the **Supplementary Materials**.

### Visualization of population structure

We used principal component analysis and t-Distributed Stochastic Neighbor Embedding (t-SNE) to summarize the population structure of the study participants to the level of the continental populations in the reference datasets (1KGP and PAGE Study) (29,30). Details on population structure estimation and visualization are provided in the **Supplementary Materials**.

### MSI status determination

MSI status of the tumors was determined based on dMMR status from clinical records when available (n=182, 25.3% of study participants). For cases with missing clinical information (n=536), MSI status was derived from tumor-normal paired whole exome sequence data using MSIsensor (version 0.5), with the percentage of mutated microsatellite loci expressed as the MSI score from 0% to 100% (31). Tumors were classified as MSI-H if MSI score >10% and MSS if MSI score ≤10%, based on a cutoff established in previous studies (32,33).

### Tumor mutational burden quantification

TMB was defined as the total number of protein-altering mutations detected in the targeted exome region and computed as total mutation counts. Using the MSI score-defined MSI status, we determined an empirical cutoff of natural log-transformed TMB at 6.2 (approx. 493 total mutations) to categorize patients into hypermutated (≤6.2, n=109) and non-hypermutated (>6.2, n=609) groups (**Supplementary Fig. 1**).

### Robust regression analysis

Somatic mutation data were analyzed through the robust regression procedure to identify potential CRC driver genes based on higher-than-expected mutation rates after controlling for gene length (34). The analysis was performed in all study participants (n=718) and exclusively among Latino participants (n=128). The robust regression method was conducted using the MASS package, and results were visualized using the ‘smoothScatter’ function in R 3.6.0 (35). The spectrum of mutations for top mutated genes was visualized using OncoPrinter and MutationMapper (https://www.cbioportal.org/visualize).

### Recurrently mutated genes

Results from previous whole-exome and targeted sequencing studies of CRC were used to identify 35 recurrently mutated genes (RMGs) for focused *a priori* specified analyses (36–39). The list and source of RMGs selected for this study are presented in **Supplementary Table 4**.

### Statistical analysis

Mutations in each individual tumor were aggregated at the gene level. For each gene, patients were labeled as “1” if carrying at least one protein-altering mutation, or “0” if carrying no protein-altering mutations. Only genes with mutations occurring in ≥3 patients were retained for downstream analyses. We used multivariate logistic regression to evaluate the exome-wide associations between genetic ancestry and gene mutation status, controlling for potential confounding factors. As the sum of ancestral components at the individual level is constrained to equal one, we performed a compositional data analysis using an additive log ratio transformation (40,41), where the individual ancestral components (i.e., AFR, EAS, NAT) were expressed in terms of additive log ratios with respect to a fixed reference component. Since EUR ancestry was the predominant ancestral component in most study samples, we used this as a reference in the log ratio transformation. The log ratios were then fit into a logistic regression model to test the overall contribution of genetic ancestry to somatic mutational status through a 3-degree of freedom likelihood-ratio test (3-df LRT). False Discovery Rate (FDR) correction was performed by the Benjamini-Hochberg procedure to adjust p-values for multiple testing, and the resulting FDR-adjusted p-values were presented (42). We also performed ancestral component-specific logistic regression models to separately estimate the odds ratio (OR) and 95% confidence interval (CI) for the associations between individual ancestral components and gene mutational status. The ancestral component was stratified into deciles and included in the model as a continuous variable to estimate the risk associated with each 10% increase in genetic ancestry. Genes were determined to be significantly associated with genetic ancestry if both the 3-df LRT FDR p-value (from the compositional data analysis) and the Wald test p-value (from the ancestral component-specific analysis) were <0.05. For RMGs, statistical significance was defined by a Wald test p-value <0.05 from both the compositional data analysis and the ancestral component-specific analysis due to pre-specified hypotheses. Statistical tests were two-sided.

Genes associated with genetic ancestry overall (3-df LRT FDR p<0.05) as well as RMGs were further tested for associations with ethnicity in multivariate logistic regression models. For each gene, we estimated the OR and 95% CI for the association with Latino versus non-Latino ethnicity. Genes were significantly associated with ethnicity if the Wald-test p-value was <0.05.

Compositional data analysis was used to test the associations between genetic ancestry with MSI status (MSI-H vs MSS; logistic regression) or TMB (linear regression). MSI status and TMB were also tested for their associations with individual ancestral components as dichotomous variables, where we used a cut-off of the mean value of the ancestral component in all study participants (≤mean vs >mean ancestry) as well as each 10% increase in genetic ancestry proportion.

To understand if hypermutation status was driving the associations between genetic ancestry and gene mutation status observed in our study, we performed a sensitivity analysis in hypermutated and non-hypermutated patients separately.

All models were adjusted for covariates selected a priori informed by clinical knowledge: age at diagnosis, sex (male, female), tumor location (colon, rectum), and tumor stage (I, II, III, IV). To maintain sufficient sample size, missing values for tumor location (n=75) and tumor stage (n=33) were recoded as an additional category for each respective variable.

Genotype file processing, QC filtering and statistical analyses were performed using R 4.2.2 (35), Plink1.9 (43), and VCFtools (44).

## Results

### Population structure and global genetic ancestry

We first characterized global genetic ancestry proportions based on genetic similarities using germline whole exome sequencing data on 718 CRC patients (128 Latino, 469 non-Latino). Principal component analysis showed that the study participants clustered with continental reference groups from the 1000 Genomes Project (1KGP) and the Population Architecture using Genomics and Epidemiology (PAGE) Study (**Fig. 1A**). Principal components (PC) 1, 2 and 3 accounted for 47.7%, 28.3% and 15.6%, respectively, of the genetic variance across the first 10 PCs, and none of the trailing PCs accounted for more than 2.0%. PC1 distinguished individuals based on AFR ancestry, while PC2 discerned EUR ancestry. Using an alternative framework (t-SNE), we confirmed that non-Latino patients were primarily concentrated within the range of variation of EUR reference population individuals, while Latino patients clustered between the EUR and NAT reference groups (**Fig. 1B**). In **Fig. 1C** and **1D**, we show the distinct ancestry patterns defining participants who identified as Latinos versus non-Latinos. EUR ancestry was the most prevalent ancestral component in both Latino and non-Latino patients; however, the average proportions of NAT and AFR ancestries were statistically significantly higher in Latino patients (20.2% and 15.2%, respectively) compared to non-Latino patients (0.5% and 10.1%, respectively; p<2.2×10^-16^, **Supplementary Table 5**). The high AFR ancestry in the Latino group was mostly observed in Puerto Ricans from the Puerto Rico Biobank (PRBB) and Black or African American TCGA patients who identified as Hispanic, while primarily Mexican-origin Latino participants from HCCS had the highest level of NAT ancestry (**Supplementary Table 6** and **Supplementary Fig. 2**).

**Figure 1.**
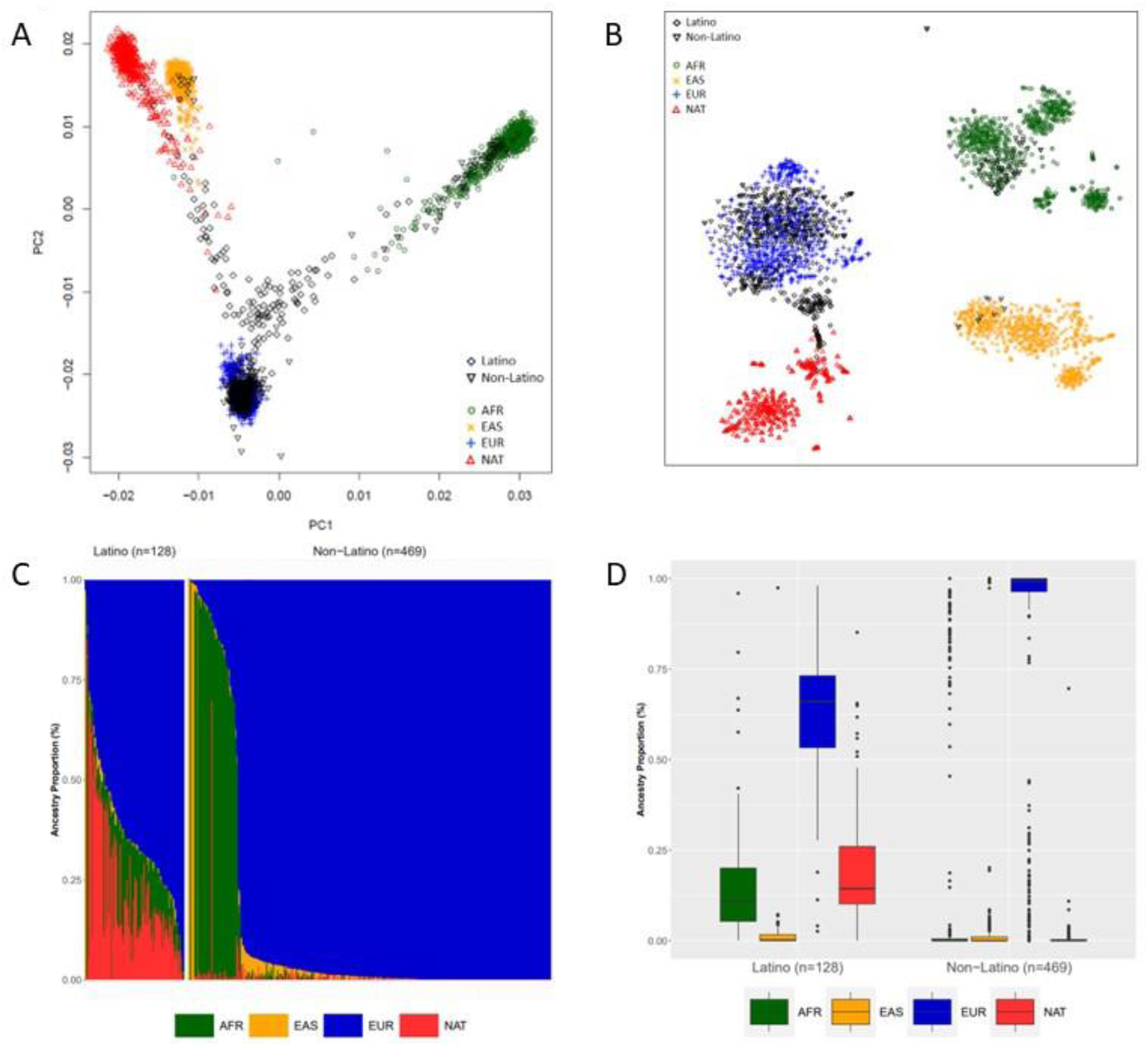
Estimated population structure and genetic ancestry for 718 colorectal cancer patients stratified by ethnicity (Latino vs non-Latino). A. Principal component analysis (PCA) for Latino and non-Latino patients with the 1KGP and the PAGE Study reference samples inferred by Eigenstrat (EUR = European, EAS = East Asian, AFR = African, NAT = Native American). PC1 and PC2 refer to the first two principal components. Each patient is represented by a point, while color represents the continental reference population (European = blue; African = dark green; East Asian = orange; Amerindian = red) and the study participants stratified by ethnicity (black). B. t-SNE plot for Latino and non-Latino patients with the 1KGP and the PAGE Study reference samples (EUR = European, EAS = East Asian, AFR = African, NAT = Native American). Each patient is represented by a point, while color represents the continental reference population (European = blue; African = dark green; East Asian = orange; Amerindian = red) and the study participants stratified by ethnicity (black). C. Genetic ancestry composition for Latino vs non-Latino patients estimated through a supervised model in Admixture assuming K=4. Each patient is represented by a column partitioned into different colors corresponding to the genetic ancestral component (European = blue; African = dark green; East Asian = orange; Amerindian = red). Patients in each ethnic group are ordered by the major ancestral component in decreasing order. D. Boxplots show the distribution of each ancestral component in Latino and non-Latino patients separately. Median ancestry value is represented as a solid line, interquartile range [IQR] as a box, and whiskers extend up to 1.5*IQR from the upper and lower quartiles. Potential outliers are depicted as solid points.

### Somatic mutational landscape

In our study of paired primary colorectal tumor tissue and normal samples from 718 patients, we observed 212,832 somatic mutations across 17,764 genes, including 200,903 singletons (mutations found in only one patient’s tumor). We found that c.1305delA (p.K435fs) in *ACVR2A* was the most common mutation (observed in tumor tissue from 82 patients). Truncating mutations, defined as frameshift insertions/deletions, stop-gain, exon start/end codon changes, and essential splice sites, accounted for 20.9% (n=44,440) of all mutations identified and were detected in 76.0% of the genes (n=13,449). The remaining mutations were annotated as nonsynonymous (single base substitution) and occurred in 97.8% of the genes (n=17,376). Using robust regression analysis to control for gene length, we identified three top mutated genes above the background rate: *APC*, *TP53*, and *KRAS* (**Fig. 2A and 2B**). *APC* had the highest number of mutations in our entire dataset (n=430 mutations; n=527 tumors) followed by *TP53* (n=174 mutations; n=388 tumors) and *KRAS* (n=29 mutations; n=291 tumors). In total, mutations in *APC*, *TP53*, or *KRAS* were detected in tumors from 623 patients (86.8%). Of these, 179 mutations were observed in at least two tumors, with the most common being *KRAS* c.G35A (p.G12D, rs121913529, n=68 tumors), c.G35T (p.G12V, rs121913529, n=65 tumors), and c.G38A (p.G13D, rs112445441, n=59 tumors).

**Figure 2.**
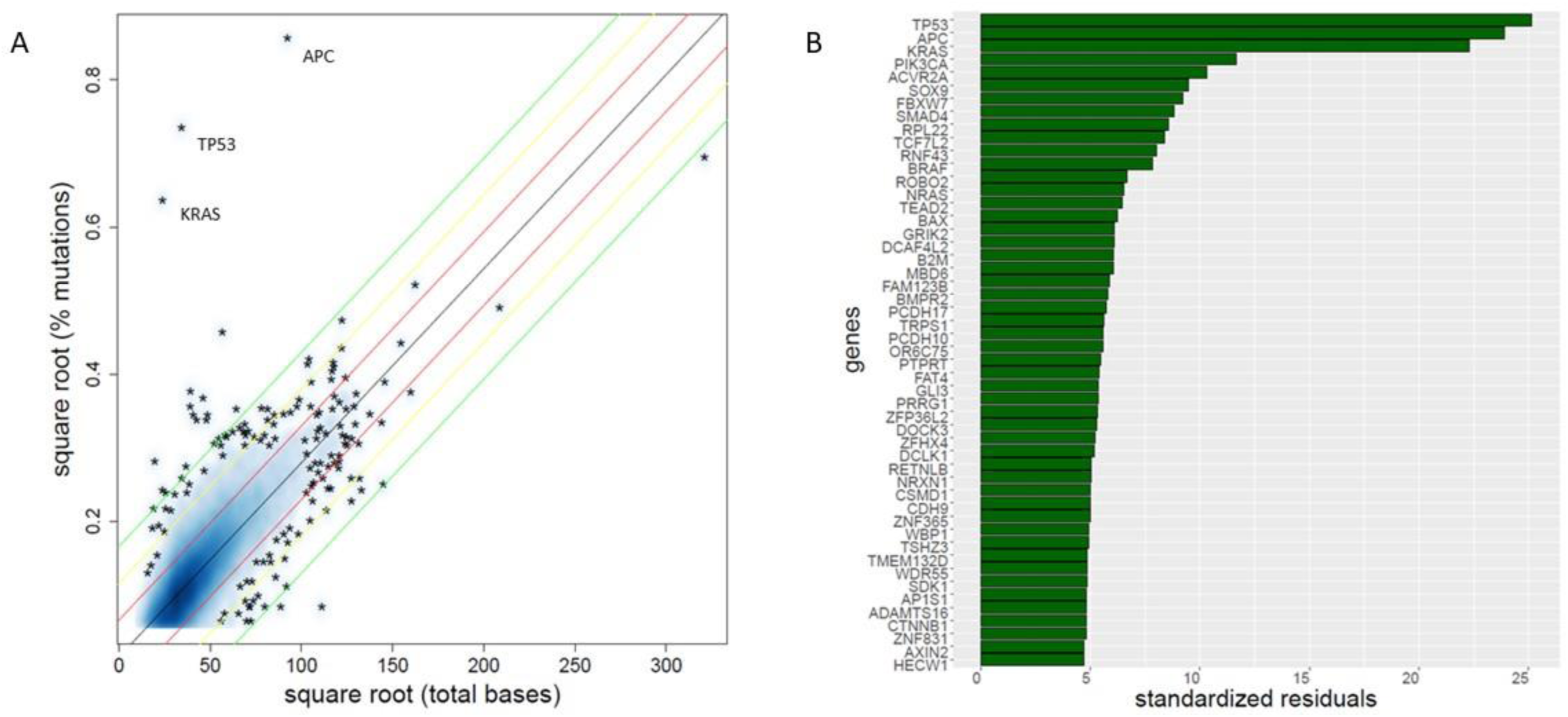
Somatic mutational landscape in 718 colorectal cancer patients. A. Percentage of mutations by total number of bases in each gene. Each point in the plot represents a gene. Genes at the top left of the plot (APC, TP53, KRAS) have a higher mutation rate relative to the gene length. B. Top 50 genes with the highest standardized residual from robust regression analysis. Each bar in the plot represents a gene. *APC*, *TP53*, and *KRAS* were the most mutated genes.

A total of 430 mutations were observed in the *APC* gene, of which 348 were protein truncating and 82 were nonsynonymous. As expected, most *APC* mutations (n=174, 40.5%) were distributed in the mutation cluster region (MCR) between codons 1200 and 1500, of which 106 resulted in a truncated APC protein. The truncating mutation c.C4348T (p.R1450X) in the MCR was the most common mutation in *APC*, found in 38 tumors (8.8%). Common truncating mutations were also found upstream of the MCR, such as c.C2626T (p.R876X, n=32 tumors) and c.C637T (p.R213X, n=24 tumors). The second most commonly over-mutated gene was *TP53* (174 mutations; 105 nonsynonymous and 69 protein truncating). Most *TP53* mutations (n=117, 67.2% of the total) were found in the DNA-binding domain (DBD) between codons 100 and 300. The DBD also harbored the most frequently mutated hotspots in *TP53*: c.G407A (p.R136H, n=49 tumors), c.C727T (p.R243W, n=28), c.G626A (p.R209Q, n=24), and c.C625T (p.R209W, n=21). *KRAS* was the third most frequently over-mutated gene with a total of 29 mutations. Except for one protein truncating mutation (G12delinsGAG), all mutations in *KRAS* were nonsynonymous mutations distributed across the Ras domain, with the most frequent mutations found in codon 12 (c.G35A, p.G12D, n=68 tumors; c.G35T, p.G12V, n=65 tumors) and codon 13 (c.G38A, p.G13D, n=59 tumors). Overall, 159 tumors had mutations in only one of these genes, while the remaining tumors had mutations in two (n=345) and three (n=119) genes. The most frequent co-mutation pattern was *APC*/*TP53* (n=197 patients) followed by *APC*/*KRAS* (n=121), *APC*/*TP53/KRAS* (n=119) and *TP53*/*KRAS* (n=27). The mutational pattern of the 50 top over-mutated genes from the robust regression analysis is illustrated in **Supplementary Fig. 3**, and lollipop charts for *APC, TP53*, and *KRAS* are presented in **Supplementary Fig. 4**.

When the analysis was restricted to tumor samples from Latino patients (n=128), *TP53*, *APC*, and *KRAS* remained the top over-mutated genes (**Supplementary Fig. 5**). Mutations in *TP53*, *APC* and *KRAS* were identified in tumors from 93 (72.7%), 67 (52.3%), and 46 (35.9%) Latino patients, respectively. In total, mutations in *APC*, *TP53*, or *KRAS* were detected in 89.1% of Latino patients (n=114).

### Association between genetic ancestry, ethnicity, and mutation status in recurrently mutated genes for CRC

We classified 35 genes as previously identified recurrently mutated genes (RMGs) in CRC based on the existing literature (see Methods; **Supplementary Table 4**) and examined their associations with genetic ancestry, after adjusting for age at diagnosis, sex, tumor location, and tumor stage (**Supplementary Table 7)**. Overall, tumors from 680 patients (94.7% of the total) harbored at least one somatic mutation in any of the RMGs. Global genetic ancestry overall was associated with *CDC27* mutation status (LRT p=0.0084), but not with individual AFR or NAT ancestry components (p>0.05). For every 10% increase in AFR ancestry, there was a 15% increase in the odds of a somatic mutation in *SMAD2* (OR=1.15, 95%CI=1.01-1.30, p=0.046). With respect to ethnicity (**Supplementary Table 8)**, Latino patients had significantly fewer *PIK3CA* mutations (13.3% vs 22.8% in non-Latino patients; OR=0.55, 95%CI=0.31-0.98, p=0.043) and *KRAS* mutations (35.9% vs 43.7% in non-Latino patients; OR=0.64, 95%CI=0.41-0.97, p=0.037). Latino patients also showed a trend toward fewer mutations in *SMAD4* (7.0%) and *TP53* (52.3%) compared to non-Latino cases (14.1% and 58.0%, respectively), but the differences did not reach statistical significance (p=0.062 and 0.074, respectively).

### Association between genetic ancestry and gene mutation status exome-wide

We next examined the associations between genetic ancestry and somatically mutated genes across the whole exome. Genetic ancestry overall was significantly associated with mutation status in 39 genes (LRT p<0.05, **Table 1**). Among these 39 genes, for each 10% increase in AFR ancestry there was a significantly higher odds of having mutations in *KNCN* (OR=1.34, 95%CI=1.09–1.66, p=5.74×10^-3^) or *TMEM184B* (OR=1.53, 95%CI=1.10–2.12, p=0.011). As shown in **Supplementary Fig. 6**, the average AFR ancestry was significantly higher in patients whose tumors had mutations in *KNCN* (39.3%) and *TMEM184B* (56.0%) relative to patients with no mutations in these genes (9.6% and 9.7%, respectively). When we repeated the analyses examining associations with ethnicity (Latino versus non-Latino), none of the 39 genes were statistically significant (**Table 2**). As shown in **Supplementary Fig. 7**, there was also no statistically significant difference between Latino and non-Latino tumors in mutation frequencies for *TMEM184B* (1.56% vs 0.21%, p=0.15) and *KNCN* (0.78% vs 1.28%, p=0.72).

**Table 1.**
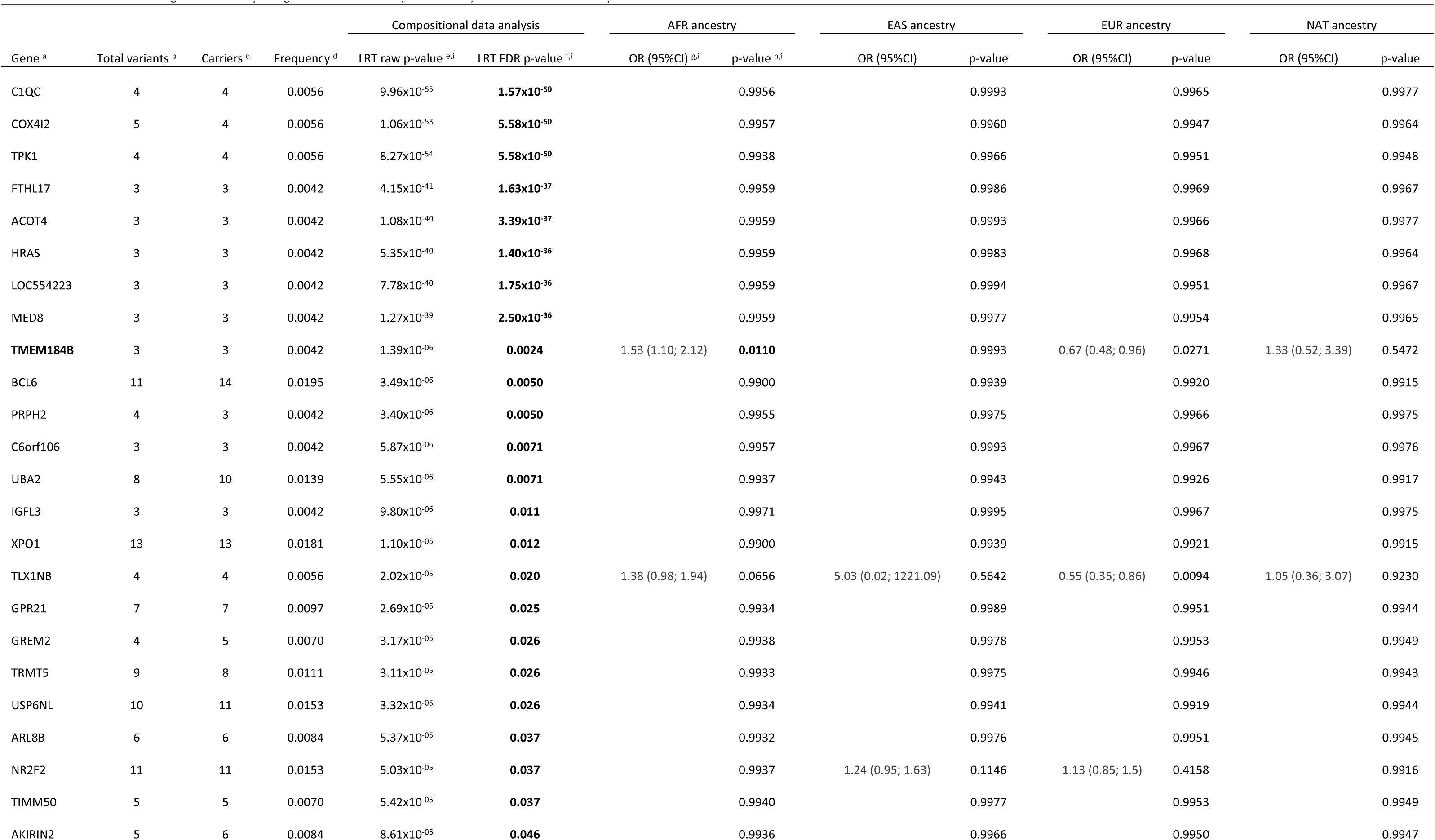
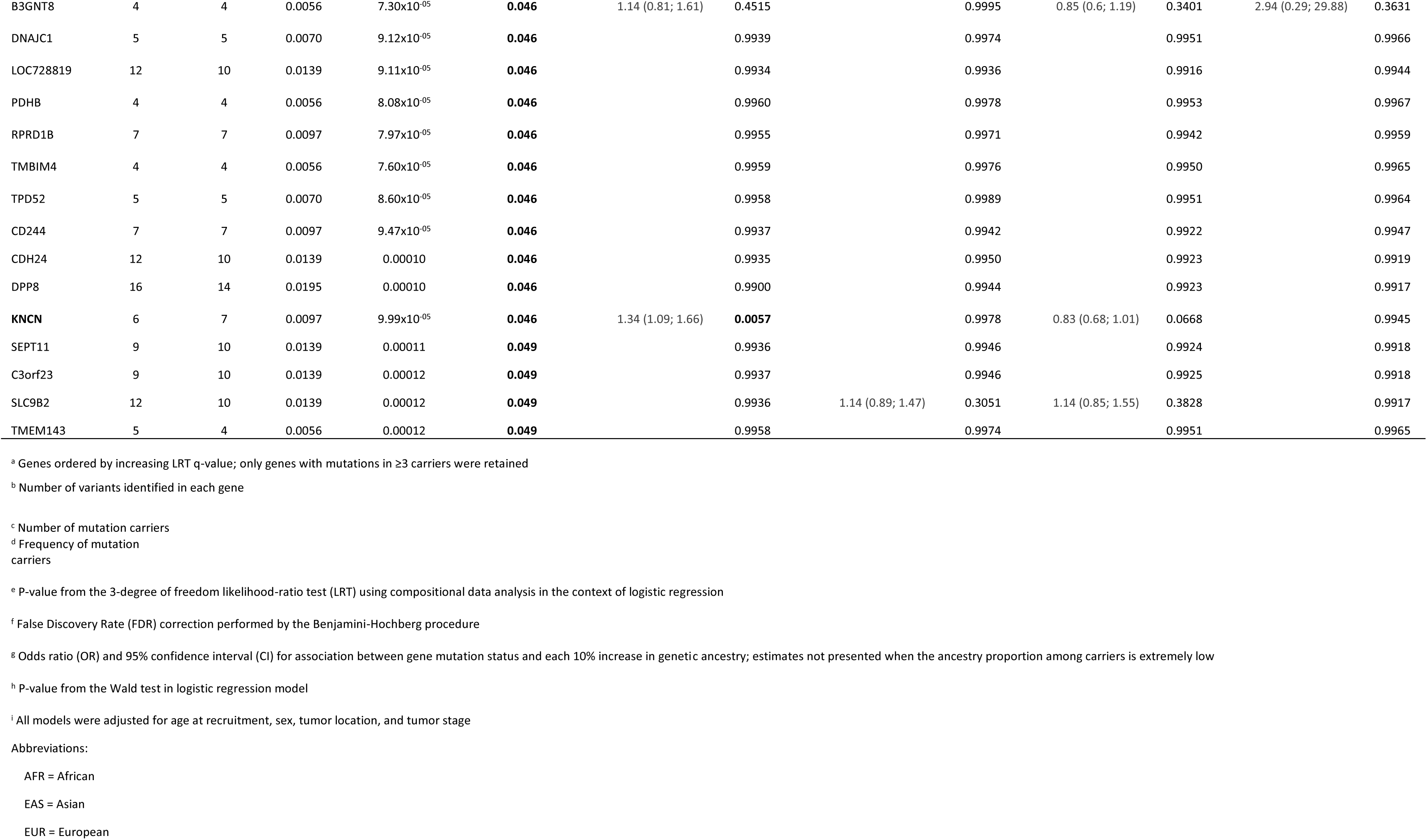
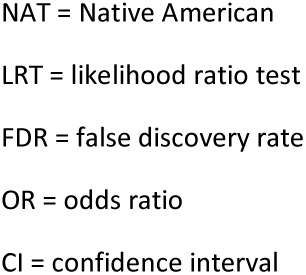
Association between genetic ancestry and gene mutation status (exome-wide) in 718 colorectal cancer patients.

**Table 2.**
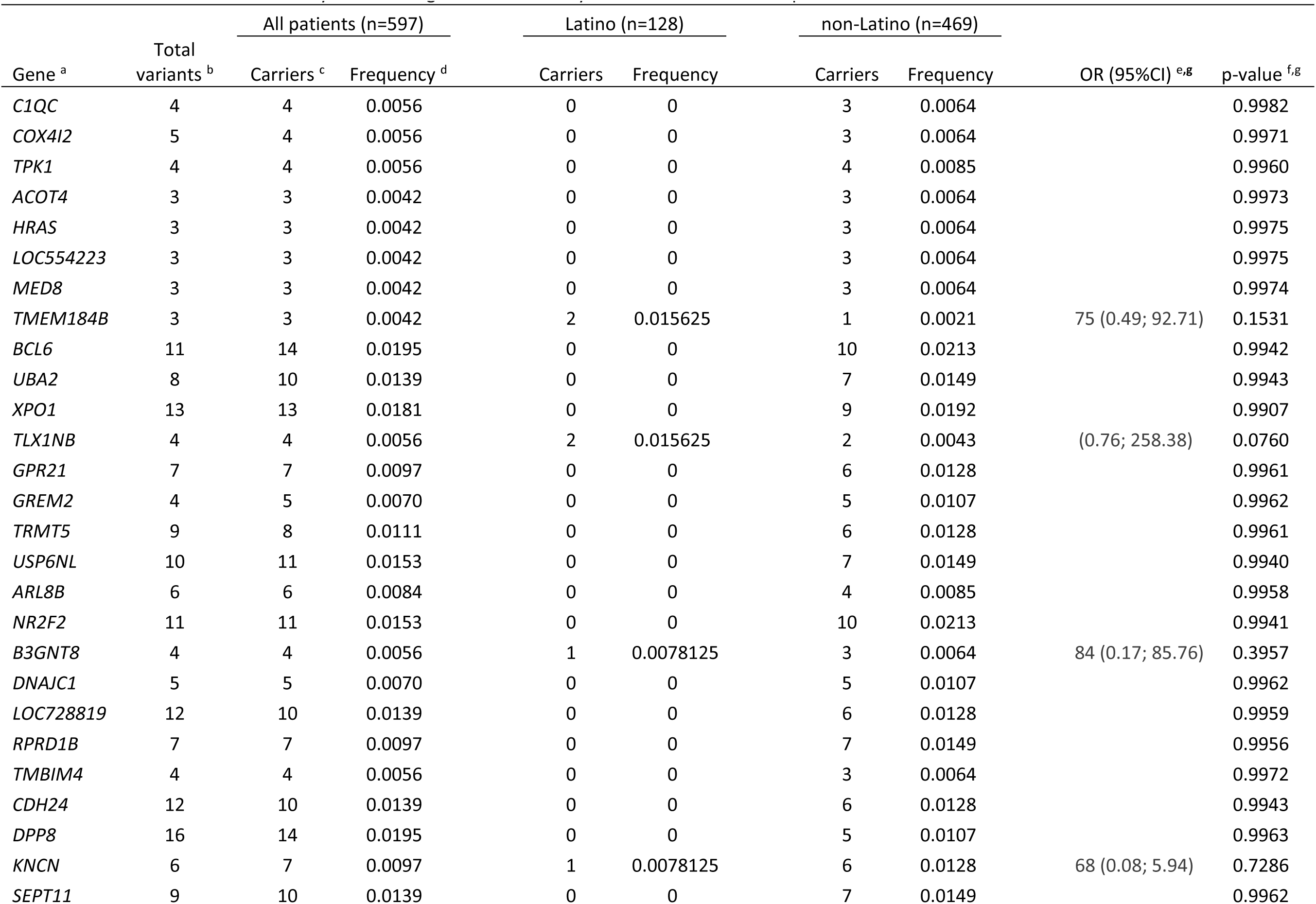
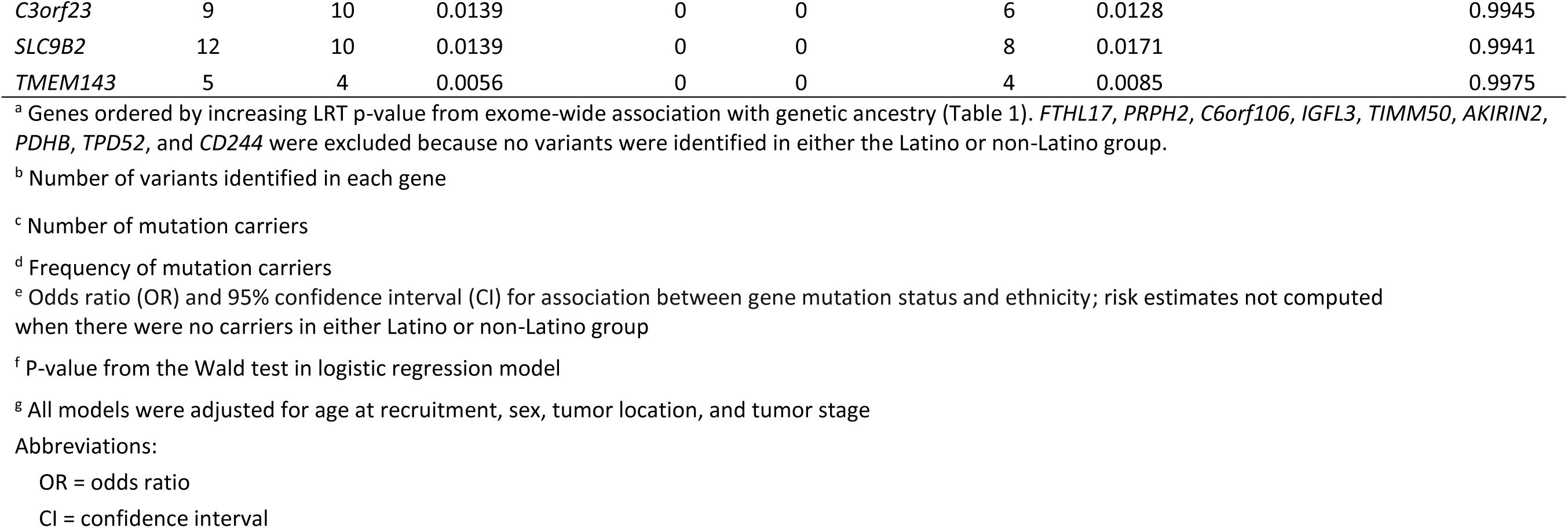
Association between ancestry-associated genes and ethnicity in 718 colorectal cancer patients.

For the ancestry-associated genes identified above, we then investigated the distribution of individual somatic mutations (**Supplementary Table 9**). For *KNCN*, a total of 6 different mutations were identified (2 protein truncating, 4 missense). All *KNCN* gene mutations occurred in the protein coding domain. Only one frameshift mutation occurred in two different patients (c.292delG, p.A98fs), while the other 5 mutations were singletons. For *TMEM184B*, we identified 3 singleton mutations: 2 missense mutations (c.C280T, p.L94F; c.G647A, p.R216H) and one protein truncating mutation (c.685delC, p.Q229fs). All three mutations were distributed in the solute transmembrane domain of the protein. Among carriers of mutations in *TMEM184B*, two patients identified as Latino and one as non-Latino.

### Associations between genetic ancestry, TMB and MSI status

Given that TMB and MSI status are molecular tumor characteristics that guide therapeutic decision making, we further examined associations between these clinical features and genetic ancestry. For MSI status, we utilized clinical data wherever available and supplemented with estimation from whole exome sequencing data (see Methods). The validity of using MSI score to classify tumors in MSI-H (MSI score >10%) and MSS (MSI score ≤10%) was confirmed by a high degree of concordance with clinical records **(Supplementary Fig.8**).

We observed a suggestive association between genetic ancestry overall and MSI status (LRT p=0.094). When genetic ancestry was dichotomized (above vs below mean value), high NAT ancestry was significantly associated with lower frequency of MSI-H tumors compared to MSS tumors (OR=0.45, 95%CI=0.21-0.99, p=0.048; **Table 3**). EAS ancestry was also inversely associated with MSI status (OR=0.31, 95%CI=0.13-0.76, p=0.01). However, there was no evidence of association with either AFR or NAT ancestry when parameterized per 10% increase (p>0.05, **Supplementary Table 10**). Genetic ancestry either as dichotomous or 10% increase was not significantly associated with TMB. In addition, TMB and MSI status were not significantly different across patients stratified by ethnicity (Latino vs non-Latino; data not shown).

**Table 3.** Association of global genetic ancestry (above vs below mean) with TMB and MSI status in 718 colorectal cancer patients.

|  | Compositional data analysis | AFR ancestry |  | EAS ancestry |  | EUR ancestry |  | NAT ancestry |  |
| --- | --- | --- | --- | --- | --- | --- | --- | --- | --- |
|  | LRT p-value <sup>a,d</sup> | OR (95%CI) <sup>b,d</sup> | p-value <sup>c,d</sup> | OR (95%CI) | p-value | OR (95%CI) | p-value | OR (95%CI) | p-value |
| TMB | 0.4702 | 0.95 (0.76; 1.18) | 0.6204 | 0.85 (0.67; 1.09) | 0.1953 | 1.05 (0.86; 1.28) | 0.6181 | 0.9 (0.72; 1.14) | 0.3853 |
| dMMR/MSI status | 0.0942 | 0.63 (0.33; 1.19) | 0.1520 | 0.31 (0.13; 0.76) | **0.0105** | 1.56 (0.89; 2.73) | 0.1212 | 0.45 (0.21; 0.99) | **0.0476** |

<sup>a</sup> P-value from the 3 degree of freedom likelihood-ratio test (LRT) using compositional data analysis in the context of logistic regression
<sup>b</sup> Odds ratio (OR) and 95% confidence interval (CI) for association between TMB or dMMR/MSI and genetic ancestry above vs below mean
<sup>c</sup> P-value from the Wald test in logistic regression model
<sup>d</sup> All models were adjusted for age at diagnosis, sex, tumor location, and tumor stage
Abbreviations:
TMB = tumor mutation burden
dMMR = deficient mismatch repair
MSI = microsatellite instability
AFR = African
EAS = Asian
EUR = European
NAT = Native American
LRT = likelihood ratio test
OR = odds ratio

### Sensitivity analysis stratified by hypermutation status

Based on TMB distribution and MSI status, we identified 109 hypermutated tumors and 609 non-hypermutated tumors (**Supplementary Fig. 1**). While tumors in the hypermutated group are usually noted as MSI-H, 14 tumors were classified as MSS. However, 10 out of these 14 patients were carriers of somatic mutations in the *POLE* exonuclease domain (P286R, V411L, S297F, P436R, A456P, F367S), which are typically associated with high mutation rate, while the remaining 4 patients had abundant frameshift mutations and relatively high MSI score (range: 5.7─9.7), suggesting some degree of MSI. In the non-hypermutated group, 8 patients’ tumors had a somatic mutation in *POLE*, but none were in the *POLE* exonuclease domain. Only one patient in the non-hypermutated group had a MSI score ≥10 (16.94) but did not carry a somatic mutation in MMR genes (*MLH1*, *MSH2*, *MSH6* and *PMS2*) or *POLE*.

We conducted sensitivity analyses stratified by hypermutation status (**Supplementary Table 11**). Among non-hypermutated patients (n=609), only 4 of the 39 genes significantly associated with genetic ancestry in the full study population carried mutations in ≥3 patients (*TMEM184B*, *RPRD1B*, *TRMT5*, *UBA2*). The mutation status of these 4 genes was significantly associated with genetic ancestry overall (LRT p<0.05), and every 10% increase in AFR ancestry was also associated with a higher odd of having mutations in *TMEM184B* (OR=1.52, 95%CI=1.09–2.11, p=0.014). Thus, where sufficient data were available, gene-level associations with genetic ancestry in the non-hypermutated group were in the same direction and had similar magnitude as in the full sample set. When the analysis was restricted to hypermutated patients (n=109), 30 out of the 39 ancestry-associated genes in the full study population had mutation in ≥3 patients. Of these, 20 genes remained statistically significantly associated with genetic ancestry overall (LRT p<0.05), including *KNCN*, while no associations with individual ancestry components were observed. A lack of observed association between *KNCN* mutation status and AFR ancestry in the hypermutated group may be driven by low sample size.

Among RMGs, mutations in *TGFBR2* were significantly associated with both genetic ancestry overall (LRT p=0.047) and each 10% increase in AFR ancestry (OR=1.40, 95%CI=1.09–1.80, p=0.009) in the hypermutated group. In the non-hypermutated group, AFR ancestry was associated with higher mutation frequency in *CTNBB1* (OR=1.15, 95%CI=1.02–1.31, p=0.027), *FAM123B* (OR=1.11, 95%CI=1.01–1.24, p=0.039) and *SMAD2* (OR=1.24, 95%CI=1.08–1.43, p=0.0025), while NAT ancestry was associated with higher mutation frequency in *CASP8* (OR=2.14, 95%CI=1.04–4.38, p=0.038); however, none of these genes were associated with genetic ancestry overall (LRT p>0.05; data not shown).

## Discussion

Here, we report findings from an exome-wide study examining the associations of genetic ancestry and Hispanic/Latino/a/x/e ethnicity with the CRC somatic mutational landscape in tumors from ancestrally diverse participants. In line with results reported for Hispanic/Latin American individuals with lung (14) and breast (15,16) cancers, we recapitulated known mutational and molecular features identified from profiling colorectal tumors from other populations, but also discovered novel associations for further follow-up studies. These results illustrate the value of examining admixed populations and assessing genetic ancestry in addition to racial and ethnic identities for informing precision medicine approaches and provide insight into genomic contributions to cancer disparities.

Consistent with data from TCGA (36), we observed that *APC*, *TP53* and *KRAS* were the most recurrently mutated genes in the overall cohort as well as in Latino patients after controlling for gene length. Our results among Latino individuals are in line with another genome sequencing analysis of 52 U.S. Latino patients with metastatic CRC where they identified commonly altered genes including *APC*, *TP53*, and *KRAS* (45). Mutations in *APC,* a well-known tumor suppressor gene, occur in approximately 80% of sporadic CRCs (36), with the MCR region being the most mutated area (46). As expected, we observed the highest frequency of mutations in the MCR region, but no differences in the prevalence of mutations by genetic ancestry or ethnicity in contrast to another report (45). For *TP53,* another widely studied tumor suppressor gene with mutations occurring in 40-50% of sporadic CRC (47), we observed a non-statistically significant lower frequency of mutations in Latino patients compared to non-Latino patients. Interestingly, *SMAD4* mutations, among the 10 top mutations in the overall cohort, were also non-significantly lower in Latino patients. Concurrent *SMAD4* and *TP53* mutations have been reported to represent a distinct poor-prognosis subgroup of mCRC (48). The proto-oncogene *KRAS* is mutated in 35%-45% of CRCs and almost all mutations are located at codons 12, 13, 61 and 146, which have been associated with resistance to therapy (49,50) and lower survival in mCRC patients (51). We also found mutations in codon 12 and codon 13 being the most frequent mutations in *KRAS*. *PIK3CA* was among the most frequently mutated genes in the overall cohort and Latino patients only. However, tumors in Latino patients had significantly fewer *KRAS* and *PIK3CA* mutations than tumors in non-Latino patients. Our findings confirm results from a previous study that reported significantly lower frequency of *PIK3CA* mutations and non-significantly lower frequency of *KRAS* mutations in Latino patients from California compared to non-Latino patients (52).

Other than non-Hispanic White populations, studies in African American patients have been the most comprehensive and offer complementary insight for interpretation of our findings. CRC patients of African descent were reported to have a younger age at diagnosis, fewer MSI-H tumors, and a significantly higher frequency of mutations in *KRAS* (including G12D/G13D), *APC*, and *PIK3CA* compared to European patients (53–55). Tumors from individuals with African ancestry also had a lower frequency of *BRAF* mutations, especially *BRAF*^V600X^ mutations, compared with European ancestry cases (53,55). No significant association between AFR ancestry and mutation frequency in any of these genes, including *BRAF*, were observed in our study. Our findings are in line with a previous study using ancestry-informative markers to identify 5,301 CRC patients of African descent, where no association between the increasing percentage of AFR ancestry and genetic alterations in cancer-related genes was found (53). Similarly, a recent study of mCRC patients reported no significant racial differences in MSI status, *KRAS*, and *BRAF* mutation rates (56). These findings suggest that genetic ancestry may not explain differences in mutation frequencies for cancer-related genes among mCRC patients. However, the proportion of AFR ancestry was only 9.9% in the overall patient population and 15.2% in Latino patients in the present study. As a result, the study cohort may lack substantial representativeness of AFR ancestry and power to identify such associations. Further studies including larger numbers of patients with a higher proportion of AFR ancestry are needed to follow-up on these observations.

On the other hand, the present study identified significant associations between AFR ancestry and increased mutation frequencies in *KNCN* and *TMEM184B*. KNCN (Kinocilin) is a cytoplasmatic protein involved in cell cycle and DNA metabolic processes (57). No studies have previously reported KNCN to be involved in carcinogenesis. TMEM184B, also known as NDC1, belongs to the transmembrane (TMEM) protein family (58) that regulates migration, proliferation and invasion through several pathways such as Mitogen-Activated Protein Kinases (MAPK), Janus Kinase/Signal Transducers, Activators of Transcription (JAK/STAT) and PhosphoInositide 3-Kinases (PI3K)/AKT (59). The TMEM family includes proteins of mostly unknown functions, but TMEMs are abnormally expressed in many malignancies (60), including CRC (61–63), and their altered expression was significantly correlated with prognosis, metastasis and drug resistance (64–66). There is also experimental evidence that TMEM proteins may act as either tumor suppressors or oncogenes (60). Further studies are needed to confirm associations with these genes and understand the underpinnings of these associations with genetic ancestry to determine if they are indicative of (i) specific loci or mutations present in specific ancestral haplotypes, or (ii) correlations between ancestral proportions and social, lifestyle and/or environmental risk factors.

Among the preselected RMGs, we found suggestive evidence that genetic ancestry is associated with mutation status in *CDC27* and *SMAD2*, which will need to be confirmed in other studies. Both genes have been implicated in carcinogenesis. *CDC27* is one of the core components of Anaphase Promoting complex/cyclosome and plays a key role in cellular division by controlling for cell cycle transitions (67). Moderate to strong expression of *CDC27* has been detected in different neoplasms, including CRC, and may play a role either like a tumor suppressor gene or oncogene (67). *SMAD2* encodes a transcriptional modulator that mediates the transforming growth factor (TGF)-beta signaling pathway, which is involved in multiple cellular processes, such as cell proliferation, apoptosis, and differentiation (68). The prevalence of *SMAD2* mutations in sporadic CRCs is 3.4% (69). Loss of *SMAD2* activation and/or expression occurs in approximately 10% of CRCs and is associated with advanced disease and poor prognosis (70).

For the NAT ancestral component, we did not observe significant differences in gene-level somatic mutations exome-wide. However, patients with high NAT ancestry had a significantly lower likelihood of having MSI-H CRC tumors compared to those with low NAT ancestry. Microsatellite instability is a biomarker of genomic alteration from an impaired DNA MMR system. CRC patients with MSI-H tumors have improved survival and response to therapy compared to same staged patients with MSS tumors (71). Previous work from our group in the Puerto Rico Biobank on 84 Puerto Rican patients, most of whom were included in the present study as well, reported that 9.4% of colon adenocarcinomas lacked expression of both MLH1 and PMS2 proteins, suggesting a lower frequency of MSI-H in the Puerto Rican population (72). An independent study of 253 CRC patients from South Florida estimated the rate of dMMR tumors in Latinos to be 12.6% and reported no significant differences in MMR deficiency by ethnicity/race (73). Lastly, a meta-analysis combining data from 22 studies reported a 12% MSI-H frequency (range, 7%-16%) in CRC tumors diagnosed among Latino patients, while MSI-H frequency was not significantly different between African Americans, Caucasians and Latinos (74). Potential differences in MSI-H frequency have implications for patient care, specifically eligibility and responsiveness to immunotherapy, and further studies are needed to clarify the relationship between genetic ancestry and MSI status, as well as the potential effect on CRC outcomes among admixed U.S. Latino patients.

The major strength of this study is the LC3 consortium infrastructure and concerted effort to characterize the somatic mutational landscapes of tumors from patients of understudied ancestral backgrounds. The larger and broader representation of Latino communities in the LC3 compared to publicly available datasets like TCGA increased the power to identify mutated genes associated with individual ancestral components and contributed to fill an important gap in our knowledge of the CRC tumor landscape of Latino individuals. In addition, the broad diversity of Native American reference samples from PAGE improved accuracy of ancestry estimation in the LC3 Latino cohort. We also used matched tumor/normal samples that minimized germline inherited variant contamination and greatly reduced the likelihood of false associations. We collected detailed clinical data on tumor characteristics and other relevant demographic, epidemiologic, and clinical variables, which allowed for adjustment based on potential confounders in multivariate models assessing the association between ancestry and tumor mutation status. We minimized multiple hypothesis testing by requiring significant genes to be associated with ancestry overall in a false discovery rate adjusted fashion before pre-specified testing for association with individual ancestry components.

There are several limitations to our study. First, as an observational study, residual confounding cannot be ruled out, and larger studies in independent samples are warranted to validate our results. Second, our current sample size was not sufficient to perform individual somatic mutation or local ancestry analysis, and these will be the focus of future work as LC3 continues to expand. While we adjusted for several potential confounders, we were limited in our ability to account for socioeconomic status, as well as certain environmental exposures, education, access to health care, and lifestyle factors which may also confound and/or modify the associations with genetic ancestry. We also did not have comprehensive information on inherited cancer syndromes such as Lynch syndrome and familial adenomatous polyposis, although we used TMB and MSI status of the tumors as a proxy for familial CRC and performed sensitivity analysis in non-hypermutated tumors only.

In summary, our results provide new knowledge relevant for precision medicine initiatives on the contribution of genetic ancestry to molecular features in CRC tumors from diverse admixed U.S. patients, including Latino individuals. Further studies are needed to elucidate the mechanisms by which genetic ancestry influences somatic mutational profiles and to evaluate the role of ancestry-associated mutations in modifying CRC outcomes.

## Data availability

Whole exome sequencing, genetic ancestry proportions and core analysis variables are available through dbGaP (phs003464).

## Supplemental material

Attached in document.

## Acknowledgements

We would like to thank all participants in the LC3 study. We would also like to thank Zhihua Chen for his analytic guidance on ancestry estimation.

## Abbreviations used in text

CRC: colorectal cancer
CI: confidence interval
HR: hazard ratio
ICD: International Classification of Diseases
LC3: Latino Colorectal Cancer Consortium
OR: odds ratio
SD: standard deviation
TCC: Total Cancer Care
HCCS: Hispanic Colorectal Cancer Study
PRBB: Puerto Rico Biobank
AFR: African
EAS: East Asian
EUR: European
NAT: Native American.

## References

1. Chakravarty D, Gao J, Phillips SM, Kundra R, Zhang H, Wang J, et al. OncoKB: A Precision Oncology Knowledge Base. JCO Precis Oncol. 2017;2017:PO.17.00011.

2. Ciardiello F, Ciardiello D, Martini G, Napolitano S, Tabernero J, Cervantes A. Clinical management of metastatic colorectal cancer in the era of precision medicine. CA Cancer J Clin. 2022;72:372– 401.

3. Zavala VA, Bracci PM, Carethers JM, Carvajal-Carmona L, Coggins NB, Cruz-Correa MR, et al. Cancer health disparities in racial/ethnic minorities in the United States. Br J Cancer. 2021;124:315–32.

4. Chu KC, Miller BA, Springfield SA. Measures of racial/ethnic health disparities in cancer mortality rates and the influence of socioeconomic status. J Natl Med Assoc. 2007;99:1092–100, 1102–4.

5. Islami F, Baeker Bispo J, Lee H, Wiese D, Yabroff KR, Bandi P, et al. American Cancer Society’s report on the status of cancer disparities in the United States, 2023. CA Cancer J Clin. 2023;

6. Alcaraz KI, Wiedt TL, Daniels EC, Yabroff KR, Guerra CE, Wender RC. Understanding and addressing social determinants to advance cancer health equity in the United States: A blueprint for practice, research, and policy. CA Cancer J Clin. 2020;70:31–46.

7. Islami F, Siegel RL, Jemal A. The changing landscape of cancer in the USA - opportunities for advancing prevention and treatment. Nat Rev Clin Oncol. 2020;17:631–49.

8. Fujimura JH, Rajagopalan R. Different differences: the use of “genetic ancestry” versus race in biomedical human genetic research. Soc Stud Sci. 2011;41:5–30.

9. Arora K, Tran TN, Kemel Y, Mehine M, Liu YL, Nandakumar S, et al. Genetic Ancestry Correlates with Somatic Differences in a Real-World Clinical Cancer Sequencing Cohort. Cancer Discov. 2022;12:2552–65.

10. Ramroop JR, Gerber MM, Toland AE. Germline Variants Impact Somatic Events during Tumorigenesis. Trends Genet. 2019;35:515–26.

11. Yuan J, Hu Z, Mahal BA, Zhao SD, Kensler KH, Pi J, et al. Integrated Analysis of Genetic Ancestry and Genomic Alterations across Cancers. Cancer Cell. 2018;34:549–560.e9.

12. Bryc K, Durand EY, Macpherson JM, Reich D, Mountain JL. The genetic ancestry of African Americans, Latinos, and European Americans across the United States. Am J Hum Genet. 2015;96:37–53.

13. Spratt DE, Chan T, Waldron L, Speers C, Feng FY, Ogunwobi OO, et al. Racial/Ethnic Disparities in Genomic Sequencing. JAMA Oncol. 2016;2:1070–4.

14. Carrot-Zhang J, Soca-Chafre G, Patterson N, Thorner AR, Nag A, Watson J, et al. Genetic Ancestry Contributes to Somatic Mutations in Lung Cancers from Admixed Latin American Populations. Cancer Discov. 2021;11:591–8.

15. Ding YC, Song H, Adamson AW, Schmolze D, Hu D, Huntsman S, et al. Profiling the Somatic Mutational Landscape of Breast Tumors from Hispanic/Latina Women Reveals Conserved and Unique Characteristics. Cancer Res. 2023;83:2600–13.

16. Romero-Cordoba SL, Salido-Guadarrama I, Rebollar-Vega R, Bautista-Piña V, Dominguez-Reyes C, Tenorio-Torres A, et al. Comprehensive omic characterization of breast cancer in Mexican-Hispanic women. Nat Commun. 2021;12:2245.

17. Schmit SL, Schumacher FR, Edlund CK, Conti DV, Ihenacho U, Wan P, et al. Genome-wide association study of colorectal cancer in Hispanics. Carcinogenesis. 2016;37:547–56.

18. Flores I, Muñoz-Antonia T, Matta J, García M, Fenstermacher D, Gutierrez S, et al. The Establishment of the First Cancer Tissue Biobank at a Hispanic-Serving Institution: A National Cancer Institute-Funded Initiative between Moffitt Cancer Center in Florida and the Ponce School of Medicine and Health Sciences in Puerto Rico. Biopreserv Biobank. 2011;9:363–71.

19. Fenstermacher DA, Wenham RM, Rollison DE, Dalton WS. Implementing personalized medicine in a cancer center. Cancer J. 2011;17:528–36.

20. Li H, Durbin R. Fast and accurate short read alignment with Burrows-Wheeler transform. Bioinformatics. 2009;25:1754–60.

21. DePristo MA, Banks E, Poplin R, Garimella KV, Maguire JR, Hartl C, et al. A framework for variation discovery and genotyping using next-generation DNA sequencing data. Nat Genet. 2011;43:491–8.

22. Saunders CT, Wong WSW, Swamy S, Becq J, Murray LJ, Cheetham RK. Strelka: accurate somatic small-variant calling from sequenced tumor-normal sample pairs. Bioinformatics. 2012;28:1811–7.

23. Cibulskis K, Lawrence MS, Carter SL, Sivachenko A, Jaffe D, Sougnez C, et al. Sensitive detection of somatic point mutations in impure and heterogeneous cancer samples. Nat Biotechnol. 2013;31:213–9.

24. Eschrich SA, Yu X, Teer JK. Fast all versus all genotype comparison using DNA/RNA sequencing data: method and workflow. BMC Bioinformatics. 2023;24:164.

25. 1000 Genomes Project Consortium, Auton A, Brooks LD, Durbin RM, Garrison EP, Kang HM, et al. A global reference for human genetic variation. Nature. 2015;526:68–74.

26. Wang K, Li M, Hakonarson H. ANNOVAR: functional annotation of genetic variants from high-throughput sequencing data. Nucleic Acids Res. 2010;38:e164.

27. Matise TC, Ambite JL, Buyske S, Carlson CS, Cole SA, Crawford DC, et al. The Next PAGE in understanding complex traits: design for the analysis of Population Architecture Using Genetics and Epidemiology (PAGE) Study. Am J Epidemiol. 2011;174:849–59.

28. Alexander DH, Novembre J, Lange K. Fast model-based estimation of ancestry in unrelated individuals. Genome Res. 2009;19:1655–64.

29. L. van der Maaten, G. Hinton. Visualizing data using t-SNE. J Mach Learn Res. 2008;9:2579–605.

30. Price AL, Patterson NJ, Plenge RM, Weinblatt ME, Shadick NA, Reich D. Principal components analysis corrects for stratification in genome-wide association studies. Nat Genet. 2006;38:904–9.

31. Niu B, Ye K, Zhang Q, Lu C, Xie M, McLellan MD, et al. MSIsensor: microsatellite instability detection using paired tumor-normal sequence data. Bioinformatics. 2014;30:1015–6.

32. Hu ZI, Shia J, Stadler ZK, Varghese AM, Capanu M, Salo-Mullen E, et al. Evaluating Mismatch Repair Deficiency in Pancreatic Adenocarcinoma: Challenges and Recommendations. Clin Cancer Res. 2018;24:1326–36.

33. Middha S, Zhang L, Nafa K, Jayakumaran G, Wong D, Kim HR, et al. Reliable Pan-Cancer Microsatellite Instability Assessment by Using Targeted Next-Generation Sequencing Data. JCO Precis Oncol. 2017;2017:PO.17.00084.

34. Schell MJ, Yang M, Teer JK, Lo FY, Madan A, Coppola D, et al. A multigene mutation classification of 468 colorectal cancers reveals a prognostic role for APC. Nat Commun. 2016;7:11743.

35. R Core Team. R: A language and environment for statistical computing. R Foundation for Statistical Computing, Vienna, Austria. 2018;

36. Cancer Genome Atlas Network. Comprehensive molecular characterization of human colon and rectal cancer. Nature. 2012;487:330–7.

37. Wood LD, Parsons DW, Jones S, Lin J, Sjöblom T, Leary RJ, et al. The genomic landscapes of human breast and colorectal cancers. Science. 2007;318:1108–13.

38. Guda K, Veigl ML, Varadan V, Nosrati A, Ravi L, Lutterbaugh J, et al. Novel recurrently mutated genes in African American colon cancers. Proc Natl Acad Sci U S A. 2015;112:1149–54.

39. Kothari N, Teer JK, Abbott AM, Srikumar T, Zhang Y, Yoder SJ, et al. Increased incidence of FBXW7 and POLE proofreading domain mutations in young adult colorectal cancers. Cancer. 2016;122:2828–35.

40. Aitchison J. The Statistical Analysis of Compositional Data. Journal of the Royal Statistical Society: Series B (Methodological). 1982;44:139–60.

41. van den Boogaart KG, Tolosana-Delgado R. Analyzing Compositional Data with R. 2013.

42. Benjamini Y, Hochberg Y. Controlling the false discovery rate: a practical and powerful approach to multiple testing. J Royal Stat Soc Ser B. 1995;57:289–300.

43. Purcell S, Neale B, Todd-Brown K, Thomas L, Ferreira MAR, Bender D, et al. PLINK: a tool set for whole-genome association and population-based linkage analyses. Am J Hum Genet. 2007;81:559– 75.

44. Danecek P, Auton A, Abecasis G, Albers CA, Banks E, DePristo MA, et al. The variant call format and VCFtools. Bioinformatics. 2011;27:2156–8.

45. Philipovskiy A, Ghafouri R, Dwivedi AK, Alvarado L, McCallum R, Maegawa F, et al. Association Between Tumor Mutation Profile and Clinical Outcomes Among Hispanic-Latino Patients With Metastatic Colorectal Cancer. Front Oncol. 2021;11:772225.

46. Miyoshi Y, Nagase H, Ando H, Horii A, Ichii S, Nakatsuru S, et al. Somatic mutations of the APC gene in colorectal tumors: mutation cluster region in the APC gene. Hum Mol Genet. 1992;1:229–33.

47. Takayama T, Miyanishi K, Hayashi T, Sato Y, Niitsu Y. Colorectal cancer: genetics of development and metastasis. J Gastroenterol. 2006;41:185–92.

48. Wang C, Sandhu J, Tsao A, Fakih M. Presence of Concurrent TP53 Mutations Is Necessary to Predict Poor Outcomes within the SMAD4 Mutated Subgroup of Metastatic Colorectal Cancer. Cancers (Basel). 2022;14:3644.

49. Dinu D, Dobre M, Panaitescu E, Bîrlă R, Iosif C, Hoara P, et al. Prognostic significance of KRAS gene mutations in colorectal cancer--preliminary study. J Med Life. 2014;7:581–7.

50. Loupakis F, Ruzzo A, Cremolini C, Vincenzi B, Salvatore L, Santini D, et al. KRAS codon 61, 146 and BRAF mutations predict resistance to cetuximab plus irinotecan in KRAS codon 12 and 13 wild-type metastatic colorectal cancer. Br J Cancer. 2009;101:715–21.

51. Alkader MS, Altaha RZ, Badwan SA, Halalmeh AI, Al-Khawaldeh MH, Atmeh MT, et al. Impact of KRAS Mutation on Survival Outcome of Patients With Metastatic Colorectal Cancer in Jordan. Cureus. 2023;15:e33736.

52. Hinduja S, Ali M, Bukari MS, Chaudhary UB, Mortenson T, Mahmood O, et al. Mutation profile of colon cancer in hispanic population of central California. JCO. 2020;38:e16067–e16067.

53. Myer PA, Lee JK, Madison RW, Pradhan K, Newberg JY, Isasi CR, et al. The Genomics of Colorectal Cancer in Populations with African and European Ancestry. Cancer Discov. 2022;12:1282–93.

54. Kang M, Shen XJ, Kim S, Araujo-Perez F, Galanko JA, Martin CF, et al. Somatic gene mutations in African Americans may predict worse outcomes in colorectal cancer. Cancer Biomark. 2013;13:359–66.

55. Jiagge E, Jin DX, Newberg JY, Perea-Chamblee T, Pekala KR, Fong C, et al. Tumor sequencing of African ancestry reveals differences in clinically relevant alterations across common cancers. Cancer Cell. 2023;41:1963–1971.e3.

56. Hinshaw TP, Fu Y, Irish WD, Parikh AA, Snyder RA. Racial Differences in Stage IV Colorectal Cancer Molecular Profiling and Mutation Rates. J Surg Res. 2023;295:763–9.

57. Leibovici M, Verpy E, Goodyear RJ, Zwaenepoel I, Blanchard S, Lainé S, et al. Initial characterization of kinocilin, a protein of the hair cell kinocilium. Hear Res. 2005;203:144–53.

58. Marx S, Dal Maso T, Chen J-W, Bury M, Wouters J, Michiels C, et al. Transmembrane (TMEM) protein family members: Poorly characterized even if essential for the metastatic process. Semin Cancer Biol. 2020;60:96–106.

59. Lemmon MA, Schlessinger J. Cell signaling by receptor tyrosine kinases. Cell. 2010;141:1117–34.

60. Schmit K, Michiels C. TMEM Proteins in Cancer: A Review. Front Pharmacol. 2018;9:1345.

61. Gao D, Han Y, Yang Y, Herman JG, Linghu E, Zhan Q, et al. Methylation of TMEM176A is an independent prognostic marker and is involved in human colorectal cancer development. Epigenetics. 2017;12:575–83.

62. Hrašovec S, Hauptman N, Glavač D, Jelenc F, Ravnik-Glavač M. TMEM25 is a candidate biomarker methylated and down-regulated in colorectal cancer. Dis Markers. 2013;34:93–104.

63. Sui Y, Sun M, Wu F, Yang L, Di W, Zhang G, et al. Inhibition of TMEM16A expression suppresses growth and invasion in human colorectal cancer cells. PLoS One. 2014;9:e115443.

64. Lin Y, Liu D, Li X, Ma Y, Pan X. TMEM184B promotes proliferation, migration and invasion, and inhibits apoptosis in hypopharyngeal squamous cell carcinoma. J Cell Mol Med. 2022;26:5551–61.

65. Qiao W, Han Y, Jin W, Tian M, Chen P, Min J, et al. Overexpression and biological function of TMEM48 in non-small cell lung carcinoma. Tumour Biol. 2016;37:2575–86.

66. Ruiz C, Martins JR, Rudin F, Schneider S, Dietsche T, Fischer CA, et al. Enhanced expression of ANO1 in head and neck squamous cell carcinoma causes cell migration and correlates with poor prognosis. PLoS One. 2012;7:e43265.

67. Kazemi-Sefat GE, Keramatipour M, Talebi S, Kavousi K, Sajed R, Kazemi-Sefat NA, et al. The importance of CDC27 in cancer: molecular pathology and clinical aspects. Cancer Cell Int. 2021;21:160.

68. Takenoshita S, Mogi A, Nagashima M, Yang K, Yagi K, Hanyu A, et al. Characterization of the MADH2/Smad2 gene, a human Mad homolog responsible for the transforming growth factor-beta and activin signal transduction pathway. Genomics. 1998;48:1–11.

69. Fleming NI, Jorissen RN, Mouradov D, Christie M, Sakthianandeswaren A, Palmieri M, et al. SMAD2, SMAD3 and SMAD4 mutations in colorectal cancer. Cancer Res. 2013;73:725–35.

70. Xie W, Rimm DL, Lin Y, Shih WJ, Reiss M. Loss of Smad signaling in human colorectal cancer is associated with advanced disease and poor prognosis. Cancer J. 2003;9:302–12.

71. Kang S, Na Y, Joung SY, Lee SI, Oh SC, Min BW. The significance of microsatellite instability in colorectal cancer after controlling for clinicopathological factors. Medicine (Baltimore). 2018;97:e0019.

72. Reverón D, López C, Gutiérrez S, Sayegh ZE, Antonia T, Dutil J, et al. Frequency of Mismatch Repair Protein Deficiency in a Puerto Rican Population with Colonic Adenoma and Adenocarcinoma. Cancer Genomics Proteomics. 2018;15:265–71.

73. Berera S, Koru-Sengul T, Miao F, Carrasquillo O, Nadji M, Zhang Y, et al. Colorectal Tumors From Different Racial and Ethnic Minorities Have Similar Rates of Mismatch Repair Deficiency. Clin Gastroenterol Hepatol. 2016;14:1163–71.

74. Ashktorab H, Ahuja S, Kannan L, Llor X, Ellis NA, Xicola RM, et al. A meta-analysis of MSI frequency and race in colorectal cancer. Oncotarget. 2016;7:34546–57.

