## Supplementary Materials for "Spectrum of somatic mutational features of colorectal tumors in ancestrally diverse populations"

*Participants and Biospecimens*

**Hispanic Colorectal Cancer Study** (HCCS). Participants in the HCCS were identified by the California Cancer Registry beginning in January 2008 (1). All men and women aged 21+ with a diagnosis of colorectal cancer (CRC) who self-identified as Hispanic or Latino and resided in California were eligible. Self-reported demographic and epidemiologic information was collected through standardized questionnaires supplemented with cancer registry data. All HCCS participants identified as Hispanic as part of the study recruitment and informed consent process. Pathology reports were used to confirm CRC diagnosis and obtain information including histology, grade, pathological stage, and tumor size. Formalin-fixed paraffin-embedded (FFPE) blocks were retrieved directly from pathology departments at the hospitals on pathology records. A total of 27 paired tumor/normal samples with available exome-wide sequencing data were included in this study. DNA was obtained from tumor tissue in FFPE blocks (tumor) and saliva (germline).

**The Puerto Rico Biobank** (PRBB). PRBB is the tissue procurement facility of the Ponce Health Sciences University-Moffitt Cancer Center Comprehensive Partnership to Advance Cancer Health Equity (2). Since 2007, self-reported Hispanic or Latino cases have been recruited from 3 sites in southern Puerto Rico (primarily San Lucas Hospital). Self-administered epidemiologic questionnaires obtained information on demographics, medical history, and lifestyle habits. A subset of PRBB samples were from study participants who either identified as Hispanic/Latino based on a study questionnaire or who were indicated as Hispanic/Latino in medical records or cancer registry data. For cases included in the PRBB through a waiver of consent, demographic data were limited, and these participants were assumed to be Hispanic/Latino. Pathology data were abstracted from hospital records and pathology reports, including anatomical site, tumor volume, TNM stage, and ulceration, among other parameters. A total of 56 paired tumor/normal DNA samples with available sequencing data were included in the present study from a combination of fresh frozen tissues, FFPE blocks (when frozen tissue was unavailable), and blood.

**Total Cancer Care Protocol (TCC)**. The TCC Protocol is the H. Lee Moffitt Cancer Center and Research Institute's on-going general biobanking protocol that began in 2006 to collect biospecimens and comprehensive epidemiologic/clinical data such as risk factors, treatment, and outcomes through linkage to electronic medical records, the Moffitt Cancer Registry, and the Florida Cancer Data System (3). At enrollment, patients were administered an electronic questionnaire to assess demographics, lifestyle factors, and medical history. Race and ethnicity information were derived from a combination of cancer registries, medical records, and electronic patient questionnaires. Pathology data were collected through medical records, pathology reports, and cancer registries. TCC contributed either frozen tumor tissue or FFPE tumor blocks and paired adjacent normal tissue or blood-derived germline DNA for these participants. Common protocols were used for tissue collection, preservation, processing, and tracking. In the current study, 216 paired tumor/normal samples with available sequencing data were included.

**The Cancer Genome Atlas (TCGA).** TCGA collected CRCs from 13 sites across Europe and the US (4). Of the 633 CRC samples available in NCI Genomic Data Commons (GDC, https://portal.gdc.cancer.gov/) as of December 2022, 314 colon (COAD) and 105 rectal (READ) cancer cases underwent WES and were integrated in the current study. The selected samples were limited to the Baylor College of Medicine (BCM) sequencing center in order to minimize batch effects. Patient and tumor information available in NCI GDC includes demographics, lifestyle factors, race, ethnicity, clinical data (e.g., stage, grade), copy number, methylation, and mRNA/miRNA expression. Biospecimen and Clinical data files were downloaded 2021-10-27 from the Genome Data Commons (https://portal.gdc.cancer.gov/repository).

*Sample selection*

All patients selected for the study were diagnosed with primary colorectal adenocarcinoma. Biospecimens from these participants that represented colorectal polyps, metastatic CRC lesions, recurrent lesions, or non-colorectal cancers were excluded. We also excluded participants that did not have paired germline/normal sequence data, had poor sequencing quality, or whose tumor and normal samples did not match by inherited variants.

*Sequence trimming*

To improve sequence alignment rates and reduce issues related to shorter template molecules, raw sequences derived from FFPE samples were trimmed to a maximum of 100 bp using cutadapt 2.10 with the following settings: "--pair-filter=any -m 30 -l 100 --trim-n -a AGATCGGAAGAGCACACGTCTGAACTCCAGTCA -A AGATCGGAAGAGCGTCGTGTAGGGAAAGAGTGT".

*Sequence alignment*

Sequences were aligned to the human reference genome version hs37d5 using the Burrows-Wheeler Aligner (BWA) (5) version 0.5.9-r16 using settings: "bwa aln -t 4 -q 15". Paired end alignment proceeded with bwa sampe: "bwa sampe -a 600". Output from bwa was converted to sorted BAM files using samtools 0.1.18 (6).

*Alignment refinement*

Duplicate reads are marked with Picard 1.56 (http://picard.sourceforge.net/). Indel realignment and Base Quality Score Recalibration (BQSR) were performed with GATK2 (GenomeAnalysisTKLite-2.2) (7). BQSR uses the following covariates: "-cov ReadGroupCovariate -cov QualityScoreCovariate -cov CycleCovariate -cov ContextCovariate".

Finally, the MD tag was added with samtools, and the BAM file was indexed. This alignment procedure was inspired by 1000 Genomes phase 2.

*Germline variant calling*

Normal (non-tumor) alignments from the study participants (n=718) were used to determine inherited variants for genetic ancestry analysis. Starting with indexed BAM files, GATK2 (GenomeAnalysisTKLite-2.2) UnifiedGenotyper was run in multi-sample mode (genotypes called on all samples at once). Variants were filtered against a common target region using BedTools v2.21.0 (8). Variant Quality Score Recalibration (VQSR) was performed with GATK2. Germline variants with genotype quality ≥15, call rate ≥95%, and all VQSR tranches except 99.90to100.00 were retained for analysis. Markers on sex chromosomes were also removed to avoid issues related to larger trait variability in heterogametic males (9). Of the 323,519 autosomal SNPs that passed quality filters, 72,508 markers overlapped by chromosome, position, reference and alternative alleles with the processed genotype data of the combined reference panel (1KGP and PAGE Study). Study participants' alleles were flipped if SNPs were genotyped on the opposite strand to match alleles in the reference panel. This set of overlapping genetic markers was used as input data for estimation of global genetic ancestry and population structure in the study participants.

*Somatic mutation detection*

Tumor and normal data files from the same individual (T/N matched pairs) were used to detect somatic mutations in the study participants (n=718). Mutations were called with both Strelka 1.0.13 (10) and MuTect 1.14 (11) (indels via GATK2Lite SomaticIndelDetector), and data were merged using custom Perl code. The GQ field in the VCF file was created to represent which methods made the call: "Best binary encoded call status: Strelka_sens=1, muTect=2, Strelka=4". Therefore, the highest quality mutations, called by both MuTect and Strelka_specific, would have a score of 6. We required a score of at least 3 (called by Strelka_specific OR (Strelka_sensitive AND MuTect) to initially consider a mutation.

*Population-level somatic mutation allele frequency filter*

To remove spurious mutations, we examined the variant allele frequency in tumor and normal samples at mutated positions. True somatic mutations would have a much higher allele frequency in tumor samples. Conversely, artifact mutations (which may be detected when allele frequency is higher in at least one tumor compared to its matched normal) would be expected to have similar allele frequencies in tumor vs normal samples at the population level. We extracted alternate allele counts from aligned BAM files from all samples at all somatic mutation positions using samtools mpileup. Alternate allele frequencies were calculated at each position, and the skewness of the allele frequency difference in tumor vs normal samples was calculated. Known driver mutations had a positive skew: tumor samples had overall much higher allele frequency at these mutations than normal samples. Artifact mutations (based on manual review of read alignments) had neutral or negative skew, indicating that the mutation frequency at these positions was similar across all tumor and normal samples. We therefore required a mutation to have a skew value ≥2 at positions with depth of coverage ≥10 in at least 80% (575/718) of the tumor and normal samples. Manual review showed that no well-known driver mutations were removed by this filter.

*Sample processing for genetic ancestry modeling*

We selected a reference dataset of 4,121 individuals from the 1,000 Genomes Project (1KGP) Phase 3 panel (n=2,504) and the Population Architecture using Genomics and Epidemiology (PAGE) Study (n=1,617) (12,13). The 1KGP data was downloaded from the International Genome Sample Resources (IGSR, www.internationalgenome.org). The 2,504 unrelated individuals extracted from the 1KGP represented 5 different continental groups: African (n=661), East Asian (n=504), European (n=503), Native American (n=347), and South Asian (n=489). The Native American and South Asian groups from the 1KGP are highly admixed (14) and were excluded. A total of 1,668 individuals were retained from the 1KGP panel. The PAGE Study from the Human Genome Diversity Project (HGDP) is a National Human Genome Research Institute (NHGRI)-created consortium representing broad diversity of Native American descent including individuals of indigenous origin from Peru, Chile, Mexico, Honduras and Colombia (http://www.pagestudy.org/). Samples from the PAGE Study were used to add representation of individual ancestral groups, especially individuals of Native American ancestry. The 1,617 unrelated individuals extracted from the PAGE Study represented the following continental groups: African (n=149), America (n=516), Central/South Asia (n=201), East Asia (n=232), Europe (n=150), Middle East (n=163), and Oceania (n=28). Individuals from highly admixed populations such as Central/South Asia, Middle East and Oceania were excluded. In addition, we excluded individuals with unknown race as well as duplicates across the 1KGP and the PAGE Study (n=178). A total of 1,047 individuals were retained from the PAGE Study.

To further remove highly admixed samples and improve accuracy in global ancestry estimation, we performed an unsupervised analysis in ADMIXTURE-1.3.0 (15) to capture the major ancestral proportions in each reference group: African (AFR), East Asian (EAS), European (EUR), and Native American (NAT). The model was initially run at various K (K=2-10), with K=4 being the minimum value that allowed to distinguish the NAT component in the study participants. A total of 872 reference individuals with the major ancestral component <98% of the total (n=410 from 1KGP; n=462 from PAGE Study) were excluded. The filtered reference datasets included 1,843 individuals (n=1,258 from 1KGP; n=585 from the PAGE Study).

Genetic data from the filtered 1KGP and PAGE Study populations were merged by chromosome, position, reference, and alternative alleles to generate a comprehensive reference panel for global ancestry estimation. Of the >81 million and 1,705,969 single nucleotide polymorphisms (SNPs) identified in 1KGP and the PAGE Study, respectively, 1,298,444 SNPs were shared between the two datasets. Monomorphic and duplicate SNPs were excluded (n=35,237). We also removed SNPs with mismatched alleles (n=83,864) but flipped alleles for SNPs genotyped on opposite strands (n=526,508) and re-formatted the data to the 1KG standard. Using duplicates across 1KGP and the PAGE Study (n=88), we computed the percentage of individuals with discordant A/T, C/G calls, before and after flipping alleles in one dataset, and retained calls with the lowest discordance rate while flipping those genotyped on opposite strands; thus, we were able to retain a large proportion of A/T, C/G calls genotyped on opposite strands (n=62,610, 48.9% of the total) that otherwise would be excluded a priori to avoid mismatching issues. We also identified and removed 9,807 SNPs with highly discordant genotype calls (discordance rate >0.2) between duplicates across the 1KGP and PAGE Study. After quality filters, 1,169,536 SNPs overlapping between the 1KGP, and the PAGE Study were merged with germline variants from the study participants and used as input data for global ancestry estimation.

*Population structure estimation*

We conducted Principal Component Analysis (PCA) as implemented by PLINK 2.0 (16) to estimate the population structure of the study participants based on continental groups in the reference individuals (1KGP and the PAGE Study). To ensure that principal components represent genome-wide structure, and not local linkage disequilibrium, the reference dataset was filtered (minor allele frequency >0.01) and pruned (plink --indep-pairwise 100 5 0.1) to remove variants with an $r^2$ value >0.1 with any other variants within a sliding window of 100 variants advanced of 5 variants at each step. LD pruning was performed separately in the Native American, African, and European reference individuals. This step generated a total of 6,712 uncorrelated SNPs in each of the 3 continental reference groups. In addition, we excluded SNPs within long-range LD regions (n=140) (17). The LD-pruned reference dataset was merged with the study subjects' genotype data. PCA was run on the combined reference and study samples using plink --pca, which returns eigenvalues and eigenvectors for the first 10 PCs.

To further characterize the continental structure of the study participants, we conducted the t-Distributed Stochastic Neighbor Embedding (t-SNE) analysis with the Rtsne package in R 3.6.0 (18,19). The 3433-sample, 72,508-SNP genotype dataset was used to create a 2-dimensional representation of the variations captured in the first 200 PCs. t-SNE was run multiple times (n=4), obtaining consistent clusters.

### References


1. Schmit SL, Schumacher FR, Edlund CK, Conti DV, Ihenacho U, Wan P, et al. Genome-wide association study of colorectal cancer in Hispanics. Carcinogenesis. 2016;37:547–56.

2. Flores I, Muñoz-Antonia T, Matta J, García M, Fenstermacher D, Gutierrez S, et al. The Establishment of the First Cancer Tissue Biobank at a Hispanic-Serving Institution: A National Cancer Institute-Funded Initiative between Moffitt Cancer Center in Florida and the Ponce School of Medicine and Health Sciences in Puerto Rico. Biopreserv Biobank. 2011;9:363–71.

3. Fenstermacher DA, Wenham RM, Rollison DE, Dalton WS. Implementing personalized medicine in a cancer center. Cancer J. 2011;17:528–36.

4. Kandoth C, McLellan MD, Vandin F, Ye K, Niu B, Lu C, et al. Mutational landscape and significance across 12 major cancer types. Nature. 2013;502:333–9.

5. Li H, Durbin R. Fast and accurate short read alignment with Burrows-Wheeler transform. Bioinformatics. 2009;25:1754–60.

6. Li H, Handsaker B, Wysoker A, Fennell T, Ruan J, Homer N, et al. The Sequence Alignment/Map format and SAMtools. Bioinformatics. 2009;25:2078–9.

7. DePristo MA, Banks E, Poplin R, Garimella KV, Maguire JR, Hartl C, et al. A framework for variation discovery and genotyping using next-generation DNA sequencing data. Nat Genet. 2011;43:491–8.

8. Quinlan AR, Hall IM. BEDTools: a flexible suite of utilities for comparing genomic features. Bioinformatics. 2010;26:841–2.

9. Reinhold K, Engqvist L. The variability is in the sex chromosomes. Evolution. 2013;67:3662–8.

10. Saunders CT, Wong WSW, Swamy S, Becq J, Murray LJ, Cheetham RK. Strelka: accurate somatic small-variant calling from sequenced tumor-normal sample pairs. Bioinformatics. 2012;28:1811–7.

11. Cibulskis K, Lawrence MS, Carter SL, Sivachenko A, Jaffe D, Sougnez C, et al. Sensitive detection of somatic point mutations in impure and heterogeneous cancer samples. Nat Biotechnol. 2013;31:213–9.

12. 1000 Genomes Project Consortium, Auton A, Brooks LD, Durbin RM, Garrison EP, Kang HM, et al. A global reference for human genetic variation. Nature. 2015;526:68–74.

13. Matise TC, Ambite JL, Buyske S, Carlson CS, Cole SA, Crawford DC, et al. The Next PAGE in understanding complex traits: design for the analysis of Population Architecture Using Genetics and Epidemiology (PAGE) Study. Am J Epidemiol. 2011;174:849–59.

14. Gravel S, Zakharia F, Moreno-Estrada A, Byrnes JK, Muzzio M, Rodriguez-Flores JL, et al. Reconstructing Native American migrations from whole-genome and whole-exome data. PLoS Genet. 2013;9:e1004023.

15. Alexander DH, Novembre J, Lange K. Fast model-based estimation of ancestry in unrelated individuals. Genome Res. 2009;19:1655–64.

16. Purcell S, Neale B, Todd-Brown K, Thomas L, Ferreira MAR, Bender D, et al. PLINK: a tool set for whole-genome association and population-based linkage analyses. Am J Hum Genet. 2007;81:559–75.

17. Price AL, Weale ME, Patterson N, Myers SR, Need AC, Shianna KV, et al. Long-range LD can confound genome scans in admixed populations. Am J Hum Genet. 2008;83:132–5; author reply 135-139.

18. R Core Team. R: A language and environment for statistical computing. R Foundation for Statistical Computing, Vienna, Austria. 2018;

19. L. van der Maaten, G. Hinton. Visualizing data using t-SNE. J Mach Learn Res. 2008;9:2579–605.
