## Supplementary Tables and Figures for "Spectrum of somatic mutational features of colorectal tumors in ancestrally diverse populations"

**Supplementary Table 1.** Demographic, clinical and tumor characteristics among 718 colorectal cancer patients

| | | Ethnicity | | |
|---|---|---|---|---|
| | All (n=718) | Latino (n=128) | Non-Latino (n=469) | Unknown (n=121) |
| **Study group [n (%)]** | | | | |
| HCCS | 27 (3.8) | 27 (21.1) | 0 (0) | 0 (0) |
| PRBB | 56 (7.8) | 56 (43.8) | 0 (0) | 0 (0) |
| TCC | 216 (30.1) | 40 (31.2) | 173 (36.9) | 3 (2.5) |
| TCGA | 419 (58.4) | 5 (3.9) | 296 (63.1) | 118 (97.5) |
| **Sex [n (%)]** | | | | |
| Female | 336 (46.8) | 51 (39.8) | 234 (42.1) | 51 (42.1) |
| Male | 382 (53.2) | 77 (60.2) | 235 (57.9) | 70 (57.9) |
| **Age at diagnosis [mean (±SD)]** | 65.2 (12.6) | 62.7 (12.9) | 64.9 (12.5) | 69.4 (11.7) |
| **Age at diagnosis [n (%)]** | | | | |
| <=50 | 94 (13.1) | 20 (15.6) | 66 (14.1) | 8 (6.6) |
| >50 | 624 (86.9) | 108 (84.4) | 403 (85.9) | 113 (93.4) |
| **Primary site [n (%)]** | | | | |
| Colon | 534 (74.4) | 90 (70.3) | 354 (75.5) | 90 (74.4) |
| Rectum | 179 (24.9) | 37 (28.9) | 111 (23.7) | 31 (25.6) |
| Unknown [a] | 5 (0.7) | 1 (0.8) | 4 (0.9) | 0 (0) |
| **Stage [n (%)]** | | | | |
| 1 | 105 (14.6) | 16 (12.5) | 59 (12.6) | 30 (24.8) |
| 2 | 241 (33.6) | 37 (28.9) | 155 (33.0) | 49 (40.5) |
| 3 | 238 (33.1) | 45 (35.2) | 169 (36.0) | 24 (19.8) |
| 4 | 98 (13.6) | 11 (8.6) | 72 (15.4) | 15 (12.4) |
| Unknown | 36 (5.0) | 19 (14.8) | 14 (3.0) | 3 (2.5) |
| **Race [n (%)]** | | | | |
| American Indian | 1 (0.1) | 0 (0) | 1 (0.2) | 0 (0) |
| Asian | 12 (1.7) | 1 (0.8) | 9 (1.9) | 2 (1.7) |
| Black | 63 (8.8) | 4 (3.1) | 55 (11.7) | 4 (3.3) |
| Other | 6 (0.8) | 5 (3.9) | 1 (0.2) | 0 (0) |
| White | 475 (66.2) | 70 (54.7) | 402 (85.7) | 3 (2.5) |
| Unknown | 161 (22.4) | 48 (37.5) | 1 (0.2) | 112 (92.6) |
| **dMMR/MSI-H status [n (%)]** [b] | | | | |
| MSI-H | 101 (14.1) | 12 (9.4) | 77 (16.4) | 12 (9.9) |
| MSS | 617 (85.9) | 116 (90.6) | 392 (83.6) | 109 (90.1) |
| **Tumor mutation burden [mean (±SD)]** [c] | 328.3 (710.6) | 233.9 (389.0) | 345.8 (723.3) | 360.0 (897.3) |
| **Hypermutation status [n (%)]** [d] | | | | |
| Non-hypermutated | 609 (84.8) | 114 (89.1) | 391 (83.4) | 104 (86.0) |

| | | | | |
|---|---|---|---|---|
| Hypermutated | 109 (15.2) | 14 (10.9) | 78 (16.6) | 17 (14.0) |

[a] Study participants with missing information

[b] For samples with missing clinical information, MSI status was determined by MSIsensor based on a MSI score cutoff of 10% (MSI-H if ≤10%; MSS if >10%)

[c] Defined as the total number of coding, non-silent mutations per patient

[d] An empirical cutoff of log-transformed mutation load at 6.6 was used to categorize samples into hypermutated(log10>6.6) and non-hypermutated (log10≤6.6)

Abbreviations:

HCCS = Hispanic Colorectal Cancer Study

PRBB = Puerto Rico Biobank

TCC = Total Cancer Care

TCGA = The Cancer Genome Atlas

**Supplementary Table 2.** Detailed sequencing analysis and quality control characteristics by sample set

| Sequencing source | Capture Kit | Sequence Provider | Sequencer | Sequence config | CRC samples (T/N pairs) |
|---|---|---|---|---|---|
| ABBV [a] | Agilent Clinical Research Exome | WashU Genomics Technology Access Center | Illumina HiSeq2500 or HiSeq3000 | 2x150 | 14 |
| ORIEN AVATAR [a] | IDT xGen + additional probes to provide double coverage on 440 actionable genes | HudsonAlpha | Illumina HiSeq4000 | 2x101 | 112 |
| BMS [a] | Agilent SureSelect v5, no UTR 51Mb kit | Expression Analysis | Illumina HiSeq | 2x101 | 90 |
| HCCS | Agilent SureSelect v6 + UTR | Macrogen (via NIH) | NovaSeq 6000 | 2x151 | 27 |
| PRBB – FF | IDT xGen Exome | Fulgent | Illumina HiSeq | 2x76 | 24 |
| PRBB – FFPE [b] | NimbleGen SeqCap EZ MedExome | Fulgent | Illumina HiSeq | 2x151 | 32 |
| TCGA | Unknown (used refseq + 25bp flank) | NA | NA | NA | 419 |

[a] TCC (Total Cancer Care) included ABBVIE, AVATAR and BMS cohorts

[b] Normal (non-tumor) samples from 3 PRBB samples were genotyped on the IDT xGen capture kit

Abbreviations:

ABBV = AbbVie Corporation

ORIEN AVATAR = Oncology Research Information Exchange Network Avatar Research Program

BMS = Bristol Myers Squibb

HCCS = Hispanic Colorectal Cancer Study

PRBB = Puerto Rico Biobank

FF = fresh frozen

FFPE = Formalin-Fixed Paraffin-Embedded

TCGA = The Cancer Genome Atlas

**Supplementary Table 3.** Summary of the reference populations from the 1KGP and the PAGE Study

| Population | Number of unrelated individuals |
|---|---|
| **1KGP** | |
| Africa | 661 |
| Americas [a] | 347 |
| East Asia | 504 |
| Europe | 503 |
| South Asia [a] | 489 |
| **PAGE Study** [b] | |
| Africa | 149 |
| America | 516 |
| Central/South Asia [a] | 201 |
| East-Asia | 232 |
| Europe | 150 |
| Middle East [a] | 163 |
| Oceania [a] | 28 |

[a] The 1KGP samples from Americas and South Asia, and the PAGE Study samples from Central/South Asia, Middle East and Oceania were excluded from the analyses

[b] 178 duplicate samples from PAGE that were also present in 1000 Genomes were excluded from the analyses

Abbreviations:

1KGP = 1000 Genomes Project

PAGE = Population Architecture using Genomics and Epidemiology

**Supplementary Table 4.** List of recurrently mutated genes in sporadic colorectal cancer

| Gene symbol | Resources [a] | PMID |
|---|---|---|
| | 1): Wood et al. Science, 2007 | 17932254 |
| | 2): The Cancer Genome Atlas Network. Nature, 2012 | 22810696 |
| | 3): Guda et al. PNAS, 2015 | 25583493 |
| | 4): Kothari et al. Cancer, 2016 | 27244218 |
| *ACVR1B* | 2) | |
| *ACVR2A* | 2) | |
| *APC* | 1); 2) | |
| *BRAF* | 2) | |
| *CASP8* | 2) | |
| *CDC27* | 2) | |
| *CTNNB1* | 2) | |
| *EDNRB* | 2) | |
| *EPHA6* | 3) | |
| *FAM123B* | 2) | |
| *FBXW7* | 1); 2); 4) | |
| *FLCN* | 3) | |
| *FZD3* | 2) | |
| *GPC6* | 2) | |
| *HTF1F* | 3) | |
| *KIAA1804* | 2) | |
| *KRAS* | 1); 2) | |
| *MAP7* | 2) | |
| *MIER3* | 2) | |
| *MSH3* | 2) | |
| *MSH6* | 2) | |
| *MYO1B* | 2) | |
| *NRAS* | 2) | |
| *PIK3CA* | 1); 2) | |
| *POLE* | 4) | |
| *PTPN12* | 2) | |
| *SMAD2* | 2) | |
| *SMAD4* | 2) | |
| *SOX9* | 2) | |
| *TCERG1* | 2) | |
| *TCF7L2* | 2) | |
| *TGFBR2* | 2) | |
| *TP53* | 1); 2) | |
| *TTN* | 2) | |
| *WASH1* | 3) | |

[a] The first four rows refer to the selected studies; each number indicates the specific study(ies) where the corresponding gene was identified

Abbreviations:

PMID = PubMed identifier

**Supplementary Table 5.** Estimated average genetic ancestry in 718 colorectal cancer patients by ethnicity

| Mean (min-max) of ancestry % [b] | All (n=718) | Ethnicity [a] | | p-value [c] |
|---|---|---|---|---|
| | | Latino (n=128) | Non-Latino (n=469) | |
| AFR | 9.9 (0-100) | 15.2 (0.001-95.9) | 10.1 (0.001-99.9) | $<2.2x10^{-16}$ |
| NAT | 4.0 (0-85.1) | 20.2 (0.001-85.1) | 0.5 (0.001-69.6) | $<2.2x10^{-16}$ |
| EAS | 2.4 (0-100) | 1.8 (0.001-97.4) | 2.5 (0.001-99.9) | 0.093 |
| EUR | 83.8 (0-100) | 62.8 (2.6-98.0) | 86.9 (0.001-99.9) | 0.11 |

[a] Samples with unknown ethnicity (n=121) were excluded

[b] Genetic ancestry was estimated trought a supervised model in ADMIXTURE-1.3.0 and assuming K=4

[c] t-test on log transformed variables

Abbreviations:

AFR = African

EAS = East Asian

EUR = European

NAT = Native American

**Supplementary Table 6.** Estimated average genetic ancestry in 718 colorectal cancer patients by contributing study

| Mean (range) of ancestry % [b] | All (n=718) | Study cohort [a] | | | |
|---|---|---|---|---|---|
| | | HCCS (n=27) | PRBB (n=56) | TCC (n=216) | TCGA (n=419) |
| EUR | 83.8 (0-100) | 50.1 (2.6-77.8) | 66.9 (27.7-88.8) | 89.5 (2.7-100) | 85.3 (0-100) |
| EAS | 2.3 (0-100) | 5.9 (0-97.4) | 0.4 (0-2.3) | 1.1 (0-97.3) | 3.0 (0-100) |
| AFR | 9.9 (0-100) | 5.1 (0-11.2) | 19.6 (2.7-66.9) | 5.3 (0-94.9) | 11.2 (0-100) |
| AMR | 4.0 (0-85.1) | 38.9 (0-65.5) | 13.2 (4.9-24.9) | 4.0 (0-85.1) | 0.4 (0-16.4) |

[a] Samples with unknown ethnicity (n=121) were included

[b] Genetic ancestry was estimated through a supervised model in ADMIXTURE-1.3.0 and assuming K=4

Abbreviations:

HCCS = Hispanic Colorectal Cancer Study

PRBB = Puerto Rico Biobank

TCC = Total Cancer Care

TCGA = The Cancer Genome Atlas

**Supplementary Table 7.** Association between genetic ancestry and recurrently mutated genes in 718 colorectal cancer patients

| | | | | Compositional data analysis | | AFR ancestry | | EAS ancestry | | EUR ancestry | | NAT ancestry | |
|---|---|---|---|---|---|---|---|---|---|---|---|---|---|
| Gene [a] | Total variants [b] | Carriers [c] | Frequency [d] | LRT raw p-value [e,i] | LRT FDR p-value [f,i] | OR (95%CI) [g,i] | p-value [h,i] | OR (95%CI) | p-value | OR (95%CI) | p-value | OR (95%CI) | p-value |
| CDC27 | 5 | 4 | 0.0056 | **0.0084** | 0.2075 | | 0.9960 | | 0.9976 | | 0.9953 | | 0.9966 |
| PTPN12 | 14 | 18 | 0.0251 | 0.0660 | 0.4011 | 0.92 (0.71; 1.19) | 0.5440 | | 0.9942 | 1.16 (0.92; 1.46) | 0.2224 | 0.73 (0.35; 1.5) | 0.3920 |
| MIER3 | 12 | 13 | 0.0181 | 0.0719 | 0.4146 | 0.6 (0.18; 2.02) | 0.4112 | | 0.9948 | 1.47 (0.82; 2.65) | 0.1961 | 0.6 (0.14; 2.59) | 0.4937 |
| TCF7L2 | 60 | 82 | 0.1142 | 0.0795 | 0.4256 | 0.93 (0.82; 1.05) | 0.2403 | 0.95 (0.75; 1.21) | 0.6809 | 1.08 (0.98; 1.19) | 0.1413 | 0.93 (0.72; 1.21) | 0.6050 |
| PIK3CA | 70 | 150 | 0.2089 | 0.1255 | 0.5036 | 1.06 (0.98; 1.14) | 0.1306 | 0.99 (0.85; 1.16) | 0.9302 | 0.98 (0.91; 1.04) | 0.4878 | 0.87 (0.67; 1.11) | 0.2641 |
| MAP7 | 17 | 16 | 0.0223 | 0.1837 | 0.5803 | 1.03 (0.85; 1.27) | 0.7424 | 1.27 (1.04; 1.55) | **0.0185** | 0.89 (0.77; 1.04) | 0.1406 | 0.9 (0.5; 1.61) | 0.7124 |
| TCERG1 | 34 | 55 | 0.0766 | 0.2072 | 0.6091 | 0.96 (0.83; 1.1) | 0.5182 | | 0.9866 | 1.1 (0.97; 1.25) | 0.1462 | 0.83 (0.54; 1.29) | 0.4132 |
| NRAS | 14 | 42 | 0.0585 | 0.2605 | 0.6649 | 1.01 (0.87; 1.17) | 0.9090 | 0.82 (0.39; 1.75) | 0.6115 | 1.02 (0.9; 1.16) | 0.7512 | 0.77 (0.48; 1.23) | 0.2762 |
| SMAD2 | 27 | 27 | 0.0376 | 0.3329 | 0.7195 | 1.14 (1; 1.3) | **0.0462** | | 0.9904 | 0.93 (0.82; 1.05) | 0.2517 | 0.8 (0.45; 1.43) | 0.4543 |
| KIAA1804 | 39 | 39 | 0.0543 | 0.3374 | 0.7232 | 0.96 (0.81; 1.13) | 0.6232 | | 0.9906 | 1.11 (0.95; 1.31) | 0.1866 | 0.54 (0.22; 1.32) | 0.1744 |
| FZD3 | 14 | 14 | 0.0195 | 0.4407 | 0.7900 | 1.13 (0.95; 1.33) | 0.1674 | | 0.9921 | 0.94 (0.79; 1.12) | 0.5052 | 0.68 (0.26; 1.76) | 0.4269 |
| SMAD4 | 72 | 85 | 0.1184 | 0.4478 | 0.7948 | 0.99 (0.9; 1.1) | 0.8908 | 1.09 (0.91; 1.3) | 0.3657 | 1 (0.92; 1.1) | 0.9444 | 0.92 (0.69; 1.22) | 0.5754 |
| APC | 430 | 527 | 0.7340 | 0.4679 | 0.8052 | 1.07 (0.98; 1.16) | 0.1170 | 0.91 (0.79; 1.04) | 0.1604 | 0.97 (0.91; 1.04) | 0.4043 | 1.04 (0.86; 1.27) | 0.6744 |
| GPC6 | 17 | 17 | 0.0237 | 0.4794 | 0.8118 | 0.9 (0.67; 1.2) | 0.4737 | | 0.9912 | 1.14 (0.89; 1.46) | 0.2966 | 0.86 (0.4; 1.84) | 0.6889 |
| ACVR2A | 28 | 102 | 0.1421 | 0.4883 | 0.8166 | 0.99 (0.9; 1.09) | 0.8080 | 1.1 (0.95; 1.27) | 0.2257 | 1 (0.92; 1.08) | 0.9955 | 0.83 (0.6; 1.15) | 0.2602 |
| FBXW7 | 72 | 97 | 0.1351 | 0.4890 | 0.8170 | 1 (0.91; 1.1) | 0.9772 | 1.03 (0.86; 1.22) | 0.7819 | 1.03 (0.94; 1.12) | 0.5133 | 0.71 (0.48; 1.04) | 0.0823 |
| FLCN | 15 | 31 | 0.0432 | 0.4951 | 0.8209 | 1.03 (0.89; 1.2) | 0.6621 | | 0.9936 | 1.02 (0.88; 1.18) | 0.8047 | 0.53 (0.16; 1.74) | 0.2955 |
| BRAF | 17 | 82 | 0.1142 | 0.5197 | 0.8357 | 0.95 (0.84; 1.06) | 0.3418 | 1.12 (0.93; 1.34) | 0.2266 | 1.05 (0.95; 1.16) | 0.3701 | 0.42 (0.17; 1.05) | 0.0630 |
| MYO1B | 21 | 18 | 0.0251 | 0.5286 | 0.8389 | 0.72 (0.39; 1.32) | 0.2840 | 1.15 (0.87; 1.51) | 0.3204 | 1.05 (0.86; 1.29) | 0.6161 | 0.97 (0.56; 1.68) | 0.9104 |
| EDNRB | 25 | 31 | 0.0432 | 0.5525 | 0.8520 | 1.01 (0.88; 1.17) | 0.8572 | | 0.9913 | 1.02 (0.89; 1.17) | 0.7989 | 0.88 (0.52; 1.49) | 0.6324 |
| TP53 | 174 | 388 | 0.5404 | 0.6221 | 0.8847 | 1.03 (0.97; 1.1) | 0.3252 | 0.94 (0.81; 1.09) | 0.3963 | 1 (0.94; 1.05) | 0.8671 | 0.94 (0.8; 1.11) | 0.4794 |
| POLE | 58 | 49 | 0.0682 | 0.6749 | 0.9062 | 0.98 (0.85; 1.12) | 0.7199 | 0.97 (0.76; 1.23) | 0.7790 | 1.03 (0.92; 1.15) | 0.6629 | 0.99 (0.72; 1.36) | 0.9675 |
| TGFBR2 | 21 | 29 | 0.0404 | 0.7045 | 0.9171 | 1.08 (0.94; 1.24) | 0.2902 | 0.99 (0.75; 1.29) | 0.9240 | 0.95 (0.83; 1.07) | 0.3896 | 1.08 (0.73; 1.58) | 0.7058 |
| KRAS | 29 | 291 | 0.4053 | 0.7188 | 0.9216 | 1.05 (0.98; 1.12) | 0.1439 | 0.94 (0.81; 1.09) | 0.3988 | 0.97 (0.92; 1.03) | 0.3778 | 1.02 (0.86; 1.2) | 0.8295 |
| FAM123B | 52 | 72 | 0.1003 | 0.7201 | 0.9216 | 1.04 (0.95; 1.15) | 0.4039 | 0.78 (0.47; 1.29) | 0.3329 | 0.99 (0.9; 1.08) | 0.7622 | 1.06 (0.8; 1.39) | 0.6841 |
| TTN | 952 | 346 | 0.4819 | 0.8250 | 0.9562 | 0.98 (0.91; 1.04) | 0.4555 | 0.99 (0.87; 1.14) | 0.9102 | 1.01 (0.96; 1.07) | 0.7048 | 1.12 (0.95; 1.32) | 0.1865 |
| MSH6 | 26 | 38 | 0.0529 | 0.8451 | 0.9600 | 1 (0.87; 1.16) | 0.9484 | 0.97 (0.74; 1.27) | 0.8237 | 0.99 (0.88; 1.12) | 0.9182 | 1.06 (0.74; 1.53) | 0.7431 |

| | | | | | | | | | | | | | |
|---|---|---|---|---|---|---|---|---|---|---|---|---|---|
| CASP8 | 27 | 28 | 0.0390 | 0.8478 | 0.9604 | 1.01 (0.86; 1.19) | 0.8725 | 1.07 (0.84; 1.37) | 0.5780 | 0.96 (0.84; 1.1) | 0.5535 | 1.15 (0.76; 1.76) | 0.5051 |
| CTNNB1 | 45 | 50 | 0.0696 | 0.8558 | 0.9617 | 1.07 (0.96; 1.19) | 0.2065 | 0.95 (0.75; 1.2) | 0.6671 | 0.97 (0.88; 1.07) | 0.4959 | 0.8 (0.52; 1.23) | 0.3124 |
| SOX9 | 87 | 91 | 0.1267 | 0.9207 | 0.9793 | 1.03 (0.94; 1.13) | 0.5672 | 1.06 (0.89; 1.27) | 0.5212 | 0.98 (0.9; 1.06) | 0.5850 | 0.96 (0.72; 1.27) | 0.7783 |
| ACVR1B | 28 | 24 | 0.0334 | 0.9314 | 0.9810 | 0.97 (0.8; 1.18) | 0.7404 | 1.22 (1.03; 1.45) | **0.0223** | 0.94 (0.82; 1.07) | 0.3627 | 0.59 (0.21; 1.65) | 0.3119 |
| MSH3 | 19 | 49 | 0.0682 | 0.9608 | 0.9886 | 1.03 (0.91; 1.16) | 0.6732 | 1.07 (0.89; 1.28) | 0.4854 | 0.95 (0.86; 1.05) | 0.3557 | 1.04 (0.75; 1.45) | 0.8101 |
| EPHA6 | 42 | 38 | 0.0529 | 0.9693 | 0.9903 | 1.01 (0.87; 1.16) | 0.9410 | | 0.9904 | 1.02 (0.9; 1.16) | 0.7537 | 1.03 (0.71; 1.47) | 0.8905 |

[a] Genes ordered by increasing LRT p-value; only genes with mutations in ≥3 carriers were retained

[b] Number of variants identified in each gene

[c] Number of mutation carriers

[d] Frequency of mutation carriers

[e] P-value from the 3-degree of freedom likelihood-ratio test (LRT) using compositional data analysis in the context of logistic regression

[f] False Discovery Rate (FDR) correction performed by the Benjamini-Hochberg procedure

[g] Odds ratio (OR) and 95% confidence interval (CI) for association between gene mutation status and each 10% increase in genetic ancestry; risk estimates not presented when the ancestry proportion among carriers is extremely low

[h] P-value from the Wald test in logistic regression model

[i] All models were adjusted for age at diagnosis, sex, tumor location, and tumor stage

Abbreviations:

AFR = African

EAS = Asian

EUR = European

NAT = Native American

LRT = likelihood ratio test

FDR = false discovery rate

OR = odds ratio

CI = confidence interval

**Supplementary Table 8.** Association between ethnicity and recurrently mutated genes in 718 colorectal cancer patients

| Gene [a] | Total variants [b] | All patients (n=597) | | Latino (n=128) | | non-Latino (n=469) | | OR (95%CI) [e,g] | p-value [f,g] |
|---|---|---|---|---|---|---|---|---|---|
| | | Carriers [c] | Frequency [d] | Carriers | Frequency | Carriers | Frequency | | |
| CDC27 | | | | | | | | | |
| PTPN12 | 14 | 18 | 0.0251 | 2 | 0.0156 | 9 | 0.0192 | 0.58 (0.11; 3.05) | 0.5198 |
| MIER3 | 12 | 13 | 0.0181 | 1 | 0.0078 | 6 | 0.0128 | 0.82 (0.1; 7.05) | 0.8555 |
| TCF7L2 | 60 | 82 | 0.1142 | 12 | 0.0938 | 57 | 0.1215 | 0.7 (0.35; 1.39) | 0.3085 |
| PIK3CA | 70 | 150 | 0.2089 | 17 | 0.1328 | 107 | 0.2281 | 0.55 (0.31; 0.98) | **0.0431** |
| MAP7 | 17 | 16 | 0.0223 | 2 | 0.0156 | 10 | 0.0213 | 0.66 (0.13; 3.3) | 0.6134 |
| TCERG1 | 34 | 55 | 0.0766 | 7 | 0.0547 | 40 | 0.0853 | 0.66 (0.28; 1.57) | 0.3506 |
| NRAS | 14 | 42 | 0.0585 | 8 | 0.0625 | 23 | 0.0490 | 1.36 (0.56; 3.27) | 0.4982 |
| SMAD2 | 27 | 27 | 0.0376 | 4 | 0.0313 | 16 | 0.0341 | 0.92 (0.29; 2.94) | 0.8854 |
| KIAA1804 | 39 | 39 | 0.0543 | 7 | 0.0547 | 26 | 0.0554 | 0.96 (0.39; 2.34) | 0.9283 |
| FZD3 | 14 | 14 | 0.0195 | 3 | 0.0234 | 7 | 0.0149 | 1.37 (0.33; 5.72) | 0.6670 |
| SMAD4 | 72 | 85 | 0.1184 | 9 | 0.0703 | 66 | 0.1407 | 0.48 (0.23; 1.04) | 0.0624 |
| APC | 430 | 527 | 0.7340 | 93 | 0.7266 | 357 | 0.7612 | 0.79 (0.5; 1.25) | 0.3110 |
| GPC6 | 17 | 17 | 0.0237 | 2 | 0.0156 | 14 | 0.0299 | 0.55 (0.12; 2.51) | 0.4421 |
| ACVR2A | 28 | 102 | 0.1421 | 14 | 0.1094 | 73 | 0.1557 | 0.72 (0.38; 1.37) | 0.3164 |
| FBXW7 | 72 | 97 | 0.1351 | 13 | 0.1016 | 67 | 0.1429 | 0.62 (0.32; 1.19) | 0.1507 |
| FLCN | 15 | 31 | 0.0432 | 5 | 0.0391 | 23 | 0.0490 | 0.83 (0.28; 2.39) | 0.7238 |
| BRAF | 17 | 82 | 0.1142 | 9 | 0.0703 | 66 | 0.1407 | 0.56 (0.26; 1.21) | 0.1406 |
| MYO1B | 21 | 18 | 0.0251 | 5 | 0.0391 | 9 | 0.0192 | 2.14 (0.66; 6.91) | 0.2025 |
| EDNRB | 25 | 31 | 0.0432 | 5 | 0.0391 | 22 | 0.0469 | 0.95 (0.35; 2.62) | 0.9254 |
| TP53 | 174 | 388 | 0.5404 | 67 | 0.5234 | 272 | 0.5800 | 0.68 (0.45; 1.04) | 0.0738 |
| POLE | 58 | 49 | 0.0682 | 8 | 0.0625 | 31 | 0.0661 | 1.01 (0.44; 2.33) | 0.9732 |
| TGFBR2 | 21 | 29 | 0.0404 | 6 | 0.0469 | 21 | 0.0448 | 0.97 (0.36; 2.64) | 0.9565 |
| KRAS | 29 | 291 | 0.4053 | 46 | 0.3594 | 205 | 0.4371 | 0.64 (0.41; 0.97) | **0.0374** |
| FAM123B | 52 | 72 | 0.1003 | 10 | 0.0781 | 50 | 0.1066 | 0.73 (0.35; 1.54) | 0.4101 |
| TTN | 952 | 346 | 0.4819 | 65 | 0.5078 | 229 | 0.4883 | 1.17 (0.78; 1.76) | 0.4454 |
| MSH6 | 26 | 38 | 0.0529 | 9 | 0.0703 | 24 | 0.0512 | 1.73 (0.75; 3.96) | 0.1976 |
| CASP8 | 27 | 28 | 0.0390 | 4 | 0.0313 | 19 | 0.0405 | 0.8 (0.26; 2.46) | 0.7022 |
| CTNNB1 | 45 | 50 | 0.0696 | 8 | 0.0625 | 39 | 0.0832 | 0.7 (0.31; 1.59) | 0.3915 |
| SOX9 | 87 | 91 | 0.1267 | 13 | 0.1016 | 67 | 0.1429 | 0.65 (0.33; 1.27) | 0.2085 |
| ACVR1B | 28 | 24 | 0.0334 | 5 | 0.0391 | 15 | 0.0320 | 1.23 (0.42; 3.63) | 0.7108 |
| MSH3 | 19 | 49 | 0.0682 | 10 | 0.0781 | 32 | 0.0682 | 1.24 (0.57; 2.67) | 0.5905 |
| EPHA6 | 42 | 38 | 0.0529 | 6 | 0.0469 | 29 | 0.0618 | 0.82 (0.33; 2.08) | 0.6785 |

[a] Genes ordered by increasing LRT p-value from exome-wide association with genetic ancestry; lines are blank for genes with no mutations in Latinos or non-Latinos

[b] Number of variants identified in each gene

[c] Number of mutation carriers

[d] Frequency of mutation carriers

[e] Odds ratio (OR) and 95% confidence interval (CI) for association between gene mutation status and ethnicity

[f] P-value from the Wald test in logistic regression model; p-values <0.05 are highlighted in bold

[g] All models were adjusted for age at diagnosis, sex, tumor location, and tumor stage

Abbreviations:

OR = odds ratio

CI = confidence interval

**Supplementary Table 9.** Characteristics of individual somatic mutations in ancestry associated genes in 718 colorectal cancer patients

| Base change | Carriers [a] | Frequency [b] | Gene | Amino acid change | Chromosome | Position | Functional effect |
|---|---|---|---|---|---|---|---|
| c.685delC | 1 | 0.0014 | TMEM184B | p.Q229fs/frameshift_deletion | 22 | 38620890 | frameshift deletion |
| c.C280T | 1 | 0.0014 | TMEM184B | p.L94F/nonsynonymous_SNV | 22 | 38626722 | nonsynonymous SNV |
| c.G647A | 1 | 0.0014 | TMEM184B | p.R216H/nonsynonymous_SNV | 22 | 38620929 | nonsynonymous SNV |
| c.G181A | 1 | 0.0014 | KNCN | p.A61T/ncRNA_intronic | 1 | 47014910 | nonsynonymous SNV |
| c.C11T | 1 | 0.0014 | KNCN | p.P4L/ncRNA_intronic | 1 | 47016877 | nonsynonymous SNV |
| c.G20A | 1 | 0.0014 | KNCN | p.S7N/ncRNA_intronic | 1 | 47016868 | nonsynonymous SNV |
| c.292delG | 2 | 0.0028 | KNCN | p.A98fs/ncRNA_intronic | 1 | 47013415 | frameshift deletion |
| c.181delG | 1 | 0.0014 | KNCN | p.A61fs/ncRNA_intronic | 1 | 47014909 | frameshift deletion |
| c.C116T | 1 | 0.0014 | KNCN | p.A39V/ncRNA_intronic | 1 | 47016772 | nonsynonymous SNV |

[a] Number of mutation carriers

[b] Frequency of mutation carriers

**Supplementary Table 10.** Association between genetic ancestry (for each 10% increase) with TMB and dMMR/MSI status in 718 colorectal cancer patients

| | Compositional data analysis | AFR ancestry | | EAS ancestry | | EUR ancestry | | NAT ancestry | |
|---|---|---|---|---|---|---|---|---|---|
| | LRT p-value [a,d] | OR (95%CI) [b,d] | p-value [c,d] | OR (95%CI) | p-value | OR (95%CI) | p-value | OR (95%CI) | p-value |
| TMB | 0.4702 | 1.01 (0.97; 1.04) | 0.7828 | 0.98 (0.9; 1.06) | 0.5711 | 1 (0.97; 1.03) | 0.9546 | 0.99 (0.91; 1.09) | 0.9169 |
| dMMR/MSI status | 0.0942 | 0.97 (0.88; 1.07) | 0.5287 | 0.99 (0.83; 1.19) | 0.9296 | 1.04 (0.96; 1.14) | 0.3394 | 0.83 (0.59; 1.18) | 0.2983 |

[a] P-value from the 3-degree of freedom likelihood-ratio test (LRT) using compositional data analysis in the context of logistic regression

[b] Odds ratio (OR) and 95% confidence interval (CI) for association between TMB or dMMR/MSI and each 10% increase of genetic ancestry

[c] P-value from the Wald test in logistic regression model

[d] All models were adjusted for age at recruitment, sex, tumor location, and tumor stage

Abbreviations:

TMB = tumor mutation burden

dMMR = deficient mismatch repair

MSI = microsatellite instability

AFR = African

EAS = Asian

EUR = European

NAT = Native American

LRT = likelihood ratio test

OR = odds ratio

CI = confidence interval

**Supplementary Table 11.** Association between genetic ancestry and gene mutation status in hypermutated and non-hypermutated patients

| | | | | Compositional data analysis | | AFR ancestry | | EAS ancestry | | EUR ancestry | | NAT ancestry | |
|---|---|---|---|---|---|---|---|---|---|---|---|---|---|
| Gene [a] | Total variants [b] | Carriers [c] | Frequency [d] | LRT raw p-value [e,i] | LRT FDR p-value [f,i] | OR (95%CI) [g,i] | p-value [h,i] | OR (95%CI) | p-value | OR (95%CI) | p-value | OR (95%CI) | p-value |
| **Non-hypermutated (n=609)** | | | | | | | | | | | | | |
| TMEM184B | 3 | 3 | 0.0049 | $2.89\times10^{-06}$ | **0.0012** | 1.52 (1.09; 2.11) | **0.0140** | | 0.9994 | 0.66 (0.46; 0.96) | 0.0283 | 1.27 (0.49; 3.27) | 0.6210 |
| UBA2 | 2 | 4 | 0.0066 | 0.0080 | 0.22 | | 0.9941 | | 0.9981 | | 0.9957 | | 0.9968 |
| TRMT5 | 4 | 3 | 0.0049 | 0.0079 | 0.22 | | 0.9961 | | 0.9986 | | 0.9952 | | 0.9967 |
| RPRD1B | 3 | 3 | 0.0049 | 0.0042 | 0.17 | | 0.9957 | | 0.9985 | | 0.9966 | | 0.9961 |
| **Hypermutated (n=109)** | | | | | | | | | | | | | |
| C1QC | 4 | 4 | 0.0367 | $9.25\times10^{-57}$ | **$1.53\times10^{-54}$** | | 0.9957 | | 1.0000 | | 0.9965 | | 0.9987 |
| COX4I2 | 4 | 3 | 0.0275 | 0.026 | 0.21 | | 0.9971 | | 0.9985 | | 0.9965 | | 0.9973 |
| FTHL17 | 3 | 3 | 0.0275 | $6.89\times10^{-28}$ | **$5.35\times10^{-26}$** | | 0.9986 | | 0.9995 | | 0.9980 | | 0.9993 |
| ACOT4 | 3 | 3 | 0.0275 | $8.44\times10^{-05}$ | **0.0042** | | 0.9980 | | 0.9991 | | 0.9968 | | 0.9986 |
| BCL6 | 11 | 14 | 0.1284 | $6.93\times10^{-07}$ | **$4.66\times10^{-05}$** | | 0.9929 | | 0.9959 | | 0.9943 | | 0.9927 |
| C6orf106 | 3 | 3 | 0.0275 | $1.18\times10^{-42}$ | **$1.32\times10^{-40}$** | | 0.9970 | | 0.9994 | | 0.9970 | | 0.9987 |
| UBA2 | 6 | 6 | 0.0550 | 0.0033 | 0.068 | | 0.9955 | | 0.9980 | | 0.9962 | | 0.9969 |
| XPO1 | 12 | 12 | 0.1101 | $2.71\times10^{-268}$ | **$5.65\times10^{-265}$** | | 0.9922 | | 0.9959 | | 0.9927 | | 0.9926 |
| TLX1NB | 3 | 3 | 0.0275 | 0.022 | 0.19 | 1.41 (0.17; 11.64) | 0.7523 | | 0.9968 | | 0.9979 | | 0.9970 |
| GPR21 | 6 | 6 | 0.0550 | $9.02\times10^{-07}$ | **$6.04\times10^{-05}$** | | 0.9955 | | 0.9989 | | 0.9946 | | 0.9979 |
| GREM2 | 4 | 5 | 0.0459 | $5.15\times10^{-08}$ | **$3.50\times10^{-06}$** | | 0.9957 | | 0.9977 | | 0.9946 | | 0.9958 |
| TRMT5 | 5 | 5 | 0.0459 | 0.0064 | 0.10 | | 0.9955 | | 0.9984 | | 0.9970 | | 0.9972 |
| USP6NL | 9 | 10 | 0.0917 | 0.00033 | **0.013** | | 0.9927 | | 0.9974 | | 0.9944 | | 0.9954 |
| ARL8B | 6 | 6 | 0.0550 | $2.12\times10^{-69}$ | **$5.08\times10^{-67}$** | | 0.9948 | | 0.9979 | | 0.9950 | | 0.9958 |
| NR2F2 | 9 | 9 | 0.0826 | $9.53\times10^{-05}$ | **0.0047** | | 0.9936 | 1.48 (1; 2.18) | 0.0504 | 1.01 (0.75; 1.36) | 0.9610 | | 0.9956 |
| TIMM50 | 5 | 5 | 0.0459 | $2.42\times10^{-71}$ | **$6.32\times10^{-69}$** | | 0.9958 | | 0.9989 | | 0.9967 | | 0.9979 |
| AKIRIN2 | 5 | 6 | 0.0550 | $8.98\times10^{-05}$ | **0.0044** | | 0.9961 | | 0.9986 | | 0.9946 | | 0.9980 |
| B3GNT8 | 3 | 3 | 0.0275 | 0.43 | 0.73 | | 0.9989 | | 0.9997 | | 0.9990 | | 0.9996 |
| DNAJC1 | 5 | 5 | 0.0459 | 0.020 | 0.18 | | 0.9960 | | 0.9989 | | 0.9967 | | 0.9986 |

| | | | | | | | | | | | | | |
|---|---|---|---|---|---|---|---|---|---|---|---|---|---|
| LOC728819 | 11 | 9 | 0.0826 | 0.00050 | **0.018** | | 0.9933 | | 0.9975 | | 0.9945 | | 0.9957 |
| RPRD1B | 4 | 4 | 0.0367 | $1.07\text{x}10^{-42}$ | **$1.23\text{x}10^{-40}$** | | 0.9975 | | 0.9996 | | 0.9979 | | 0.9983 |
| TPD52 | 5 | 5 | 0.0459 | $6.43\text{x}10^{-71}$ | **$1.65\text{x}10^{-68}$** | | 0.9957 | | 0.9991 | | 0.9965 | | 0.9978 |
| CD244 | 6 | 6 | 0.0550 | 0.0015 | **0.042** | | 0.9955 | | 0.9977 | | 0.9944 | | 0.9956 |
| CDH24 | 12 | 10 | 0.0917 | 0.0054 | 0.092 | | 0.9933 | | 0.9980 | | 0.9945 | | 0.9955 |
| DPP8 | 14 | 12 | 0.1101 | 0.0024 | 0.057 | | 0.9931 | | 0.9963 | | 0.9944 | | 0.9930 |
| KNCN | 4 | 5 | 0.0459 | 0.00026 | **0.011** | 1.62 (1.18; 2.23) | 0.0029 | | 0.9984 | 0.74 (0.57; 0.95) | 0.0195 | | 0.9958 |
| SEPT11 | 8 | 9 | 0.0826 | 0.0049 | 0.086 | | 0.9932 | | 0.9982 | | 0.9946 | | 0.9968 |
| C3orf23 | 8 | 8 | 0.0734 | 0.011 | 0.14 | | 0.9932 | | 0.9964 | | 0.9946 | | 0.9934 |
| SLC9B2 | 11 | 9 | 0.0826 | 0.00040 | **0.015** | | 0.9935 | 1.25 (0.89; 1.76) | 0.1940 | 1.05 (0.78; 1.4) | 0.7500 | | 0.9968 |
| TMEM143 | 4 | 3 | 0.0275 | $1.25\text{x}10^{-41}$ | **$1.09\text{x}10^{-39}$** | | 0.9959 | | 1.0000 | | 0.9967 | | 0.9972 |

[a] Genes ordered by increasing LRT p-value from exome-wide association with genetic ancestry (Table 1); only genes with mutations in ≥3 carriers were retained

[b] Number of variants identified in each gene

[c] Number of mutation carriers

[d] Frequency of mutation carriers

[e] P-value from the 3-degree of freedom likelihood-ratio test (LRT) using compositional data analysis in the context of logistic regression

[f] False Discovery Rate (FDR) correction performed by the Benjamini-Hochberg procedure

[g] Odds ratio (OR) and 95% confidence interval (CI) for association between gene mutation status and each 10% increase in genetic ancestry; risk estimates not presented when the ancestry proportion among carriers is extremely low

[h] P-value from the Wald test in logistic regression model

[i] All models were adjusted for age at diagnosis, sex, tumor location, and tumor stage

Abbreviations:

AFR = African

EAS = Asian

EUR = European

NAT = Native American

LRT = likelihood ratio test

FDR = false discovery rate

OR = odds ratio

CI = confidence interval

**Supplementary Figure 1**

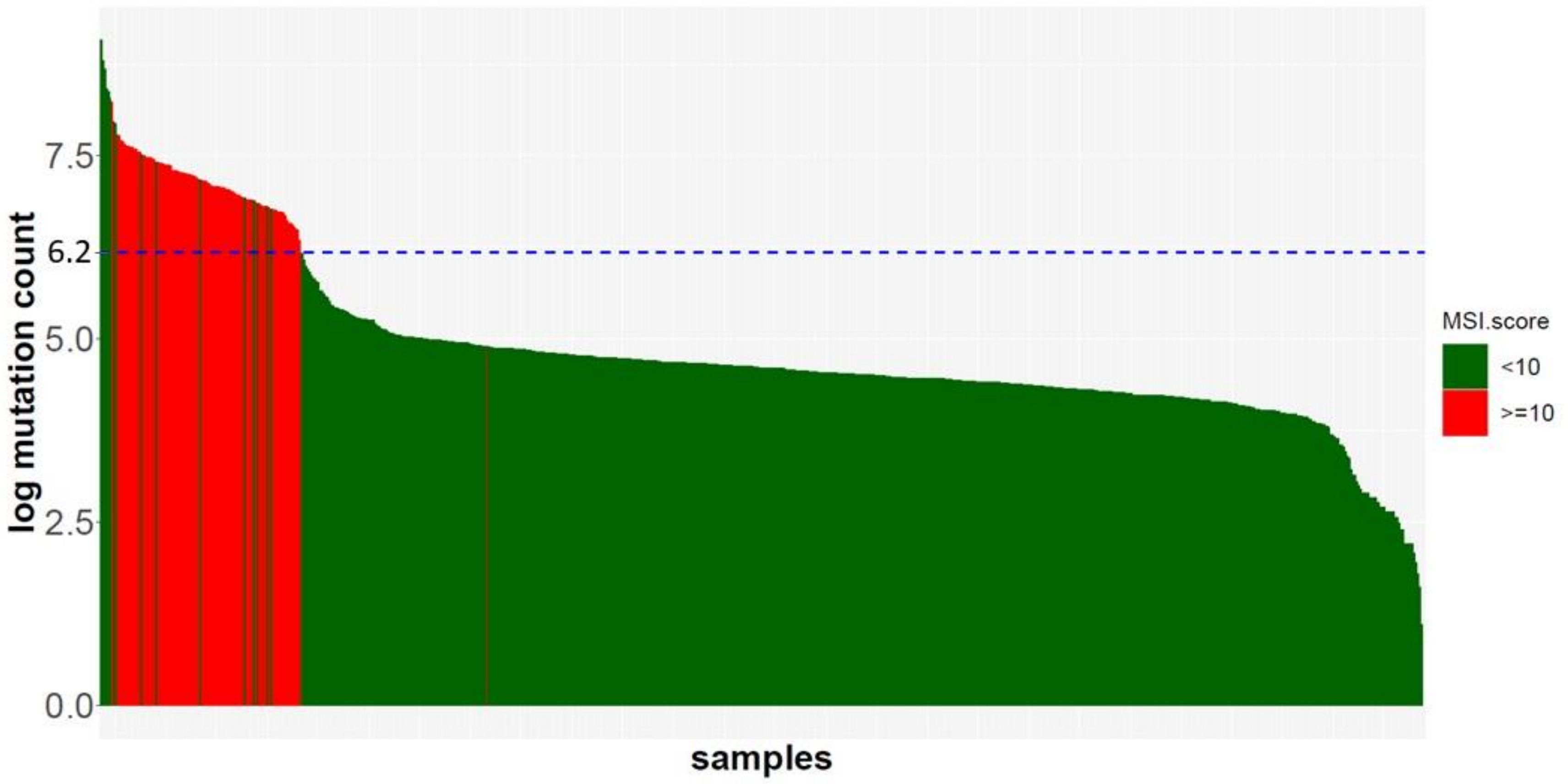


**Supplementary Figure 1.** Tumor mutational burden distribution in 718 colorectal cancer patients.
Bar colors indicate MSI score <10 (green) or ≥10 (red) for each patient. Using the dMMR/MSI status based on the MSI score, we defined an empirical cutoff of log-transformed TMB at 6.2 (blue dotted line) to categorize patients into hypermutated (≤6.2, n=109) and non-hypermutated (>6.2, n=609) groups.

**Supplementary Figure 2**

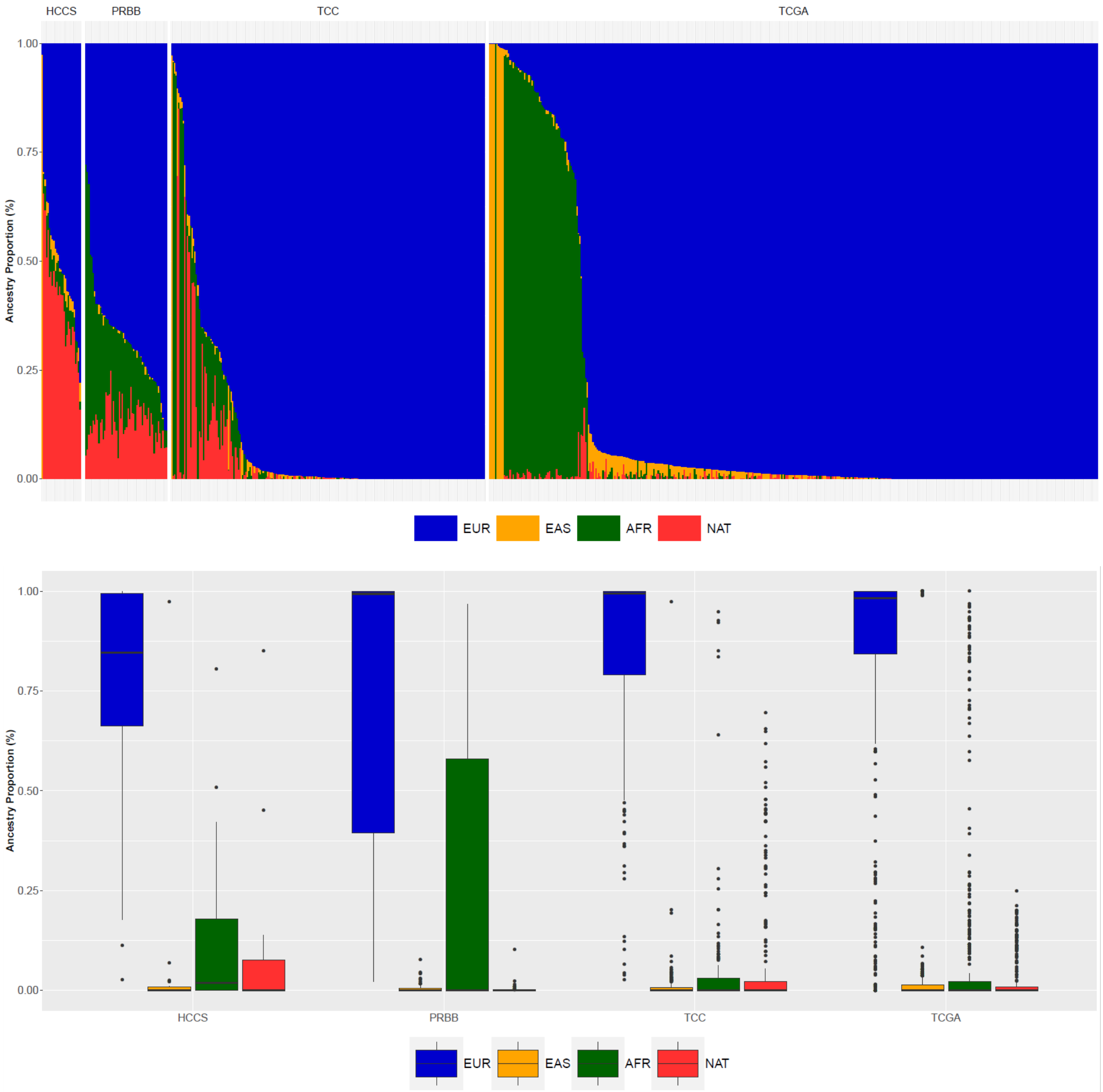


**Supplementary Figure 2.** Estimated genetic ancestry for 718 colorectal cancer patients stratified by study cohort

A. Genetic ancestry composition for Latino vs non-Latino patients estimated trought a supervised model in Admixture assuming K=4. Each patient is represented by a column partitioned into different colors corresponding to the genetic ancestral component (European = blue; African = dark green; East Asian = orange; Amerindian = red). Patients in each ethnic group are ordered by the major ancestral component in decreasing order.

B. Boxplots show the distribution of each ancestral component in Latino and non-Latino patients separately. Median ancestry value is represented as a solid line, interquartile range [IQR] as a box, and whiskers extend up to 1.5*IQR from the upper and lower quartiles. Potential outliers are depicted as solid points.

### Supplementary Figure 4

APC

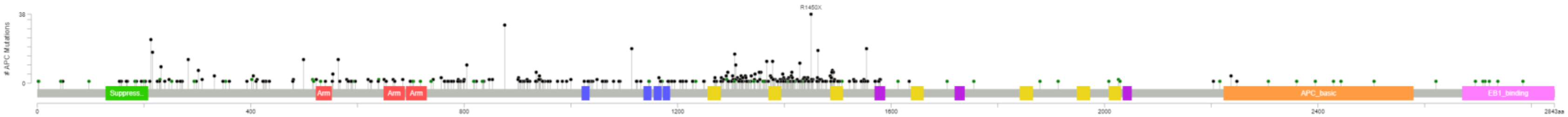

TP53

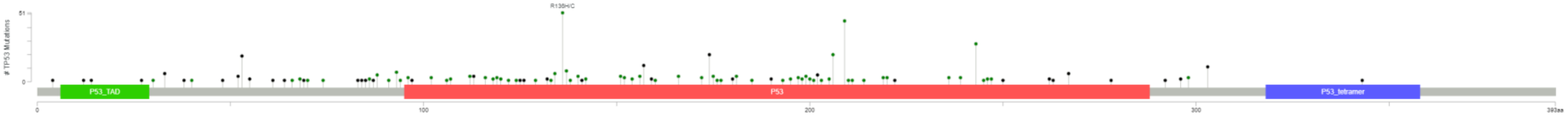

KRAS

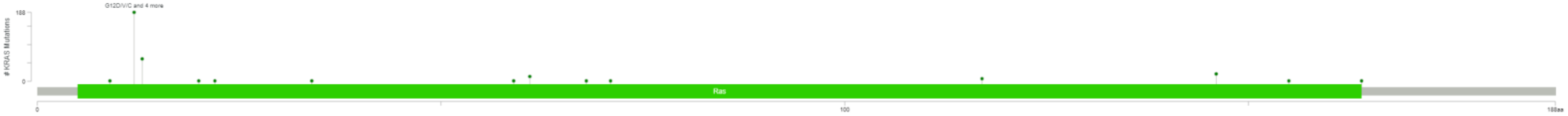

**Supplementary Figure 4.** Mutation spectrum of *APC*, *TP53*, and *KRAS* in 718 colorectal cancer patients

Lollipop-style mutation diagrams show the distribution of somatic mutation across genes. Only mutations with protein change predictions are presented. Each mutation consists of a vertical line with a dot at the higher end. Black dots correspond to truncating mutations, while green dots correspond to nonsynonymous mutations. X-axis: codon position; Y-axis: number of mutation carriers. Colored boxes represent protein domains/motifs.

**Supplementary Figure 5**

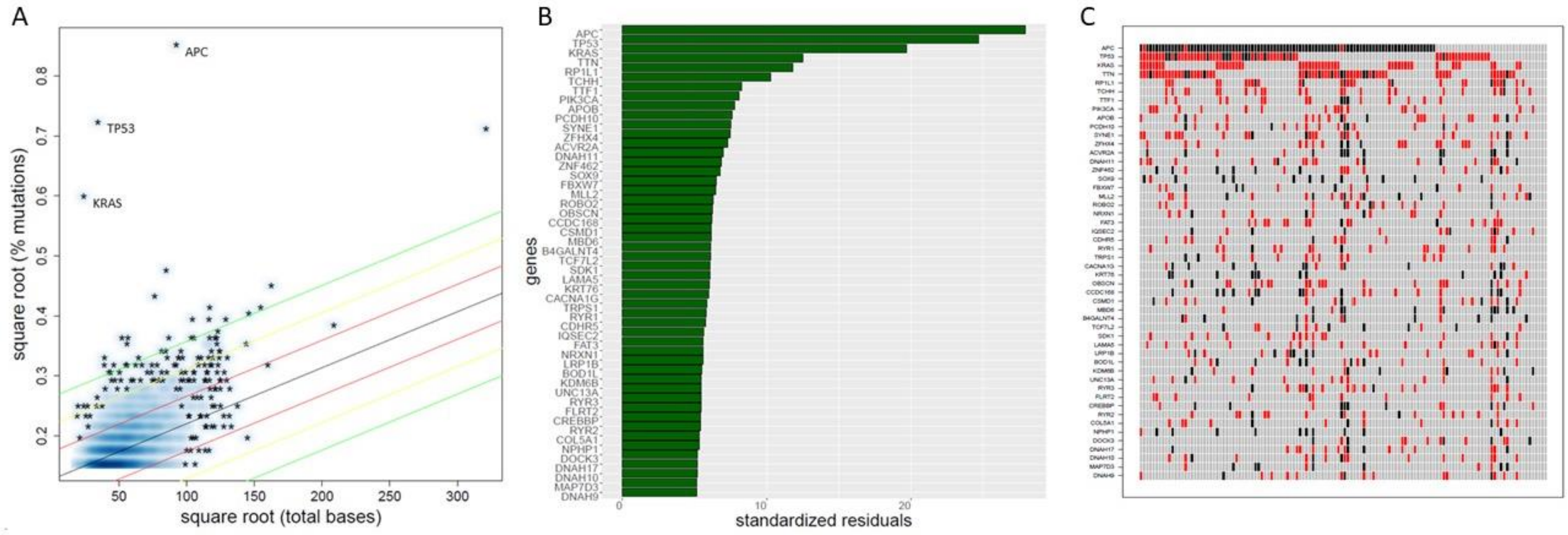


**Supplementary Figure 5.** Mutational landscape in 128 Latino colorectal cancer patients
A. Percentage of mutations by total number of bases in each gene. Each point in the plot represents a gene. Genes at the top left of the plot (APC, TP53, KRAS) have a higher mutation rate relative to the gene length.
B. Top 50 genes with the highest standardized residual from robust regression analysis. Each bar in the plot represents a gene. *APC*, *TP53*, and *KRAS* were the most mutated genes.
C. Oncoprint of the top mutated genes from robust regression analysis. Each column represents an individual patient. Red=missense mutation; Black=protein truncating mutation.

**Supplementary Figure 6**

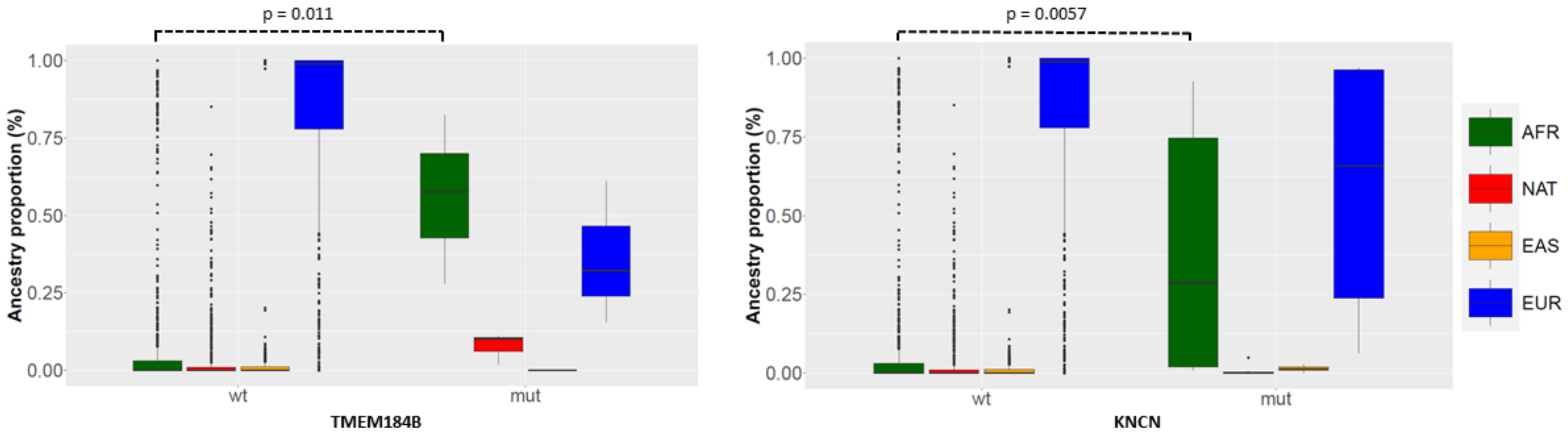


**Supplementary Figure 6.** Correlation between genetic ancestry and gene mutation status in 718 colorectal cancer patients. Average proportions of AFR (dark green), NAT (red), EAS (orange) and EUR (blue) ancestries are shown for genes with wild-type (wt) and mutated (mut) status. Boxplot lines reflect lower quartile, median, and upper quartile of ancestry, with extended points representing outliers. X-axis: mutation status (wild-type, mutated); Y-axis: genetic ancestry proportions ranging from 0 to 1. P-values from the Wald test in standard logistic regression model.

**Supplementary Figure 7**

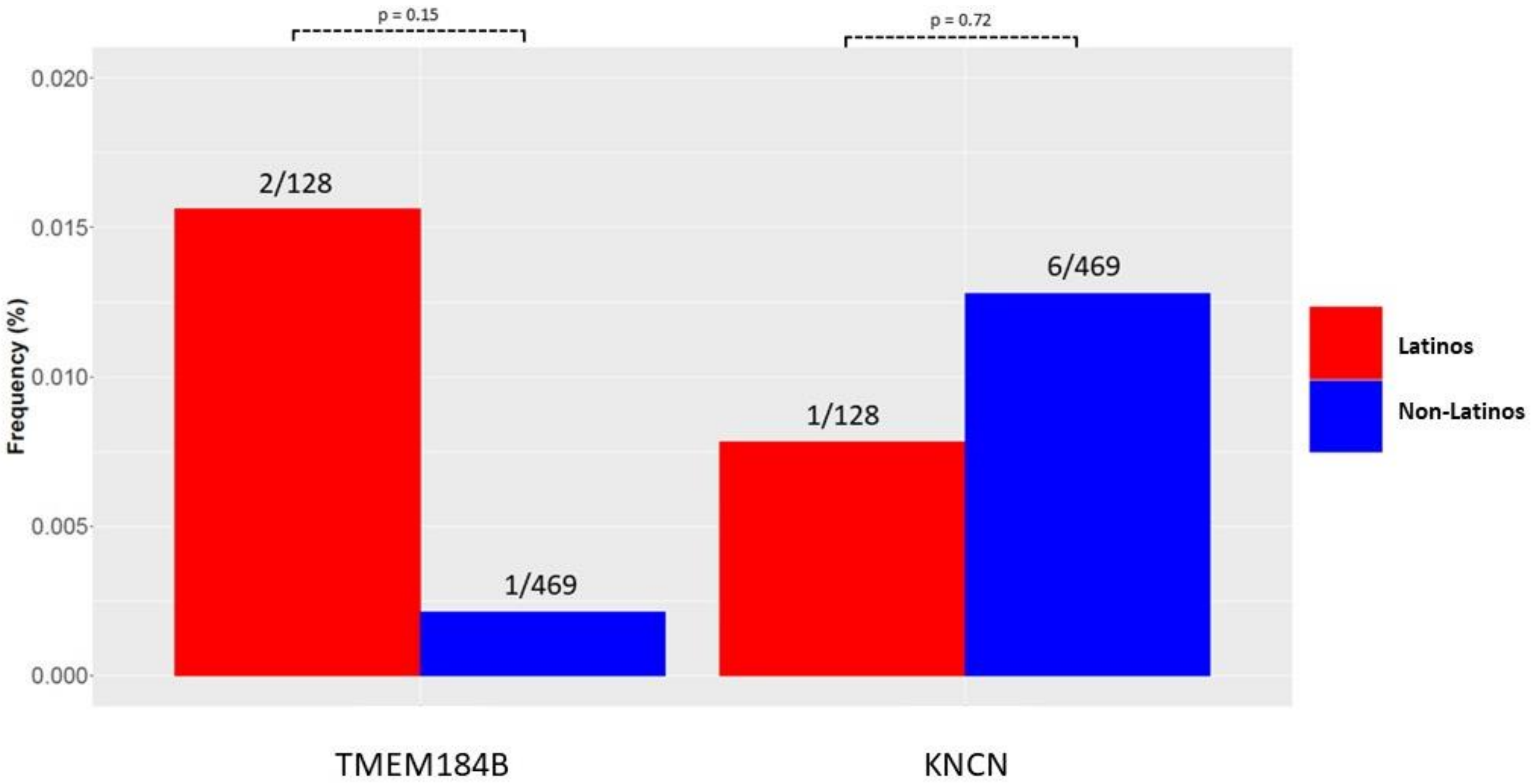


**Supplementary Figure 7.** Mutation frequency for TMEM184B and KNCN in 718 colorectal cancer patients by ethnicity. Histograms represent the proportion of patients with at least one mutation in the genes. Differences in proportions between Latinos and non-Latinos patients were tested using the Wald test from logistic regression model. P-values at different levels of statistical significance are indicated above histograms: * $p<0.05$, ** $p<0.01$, *** $p<0.001$. Mutation rates in TMEM184B and KNCN reveal no significant difference between Latino and non-Latino patients.

**Supplementary Figure 8**

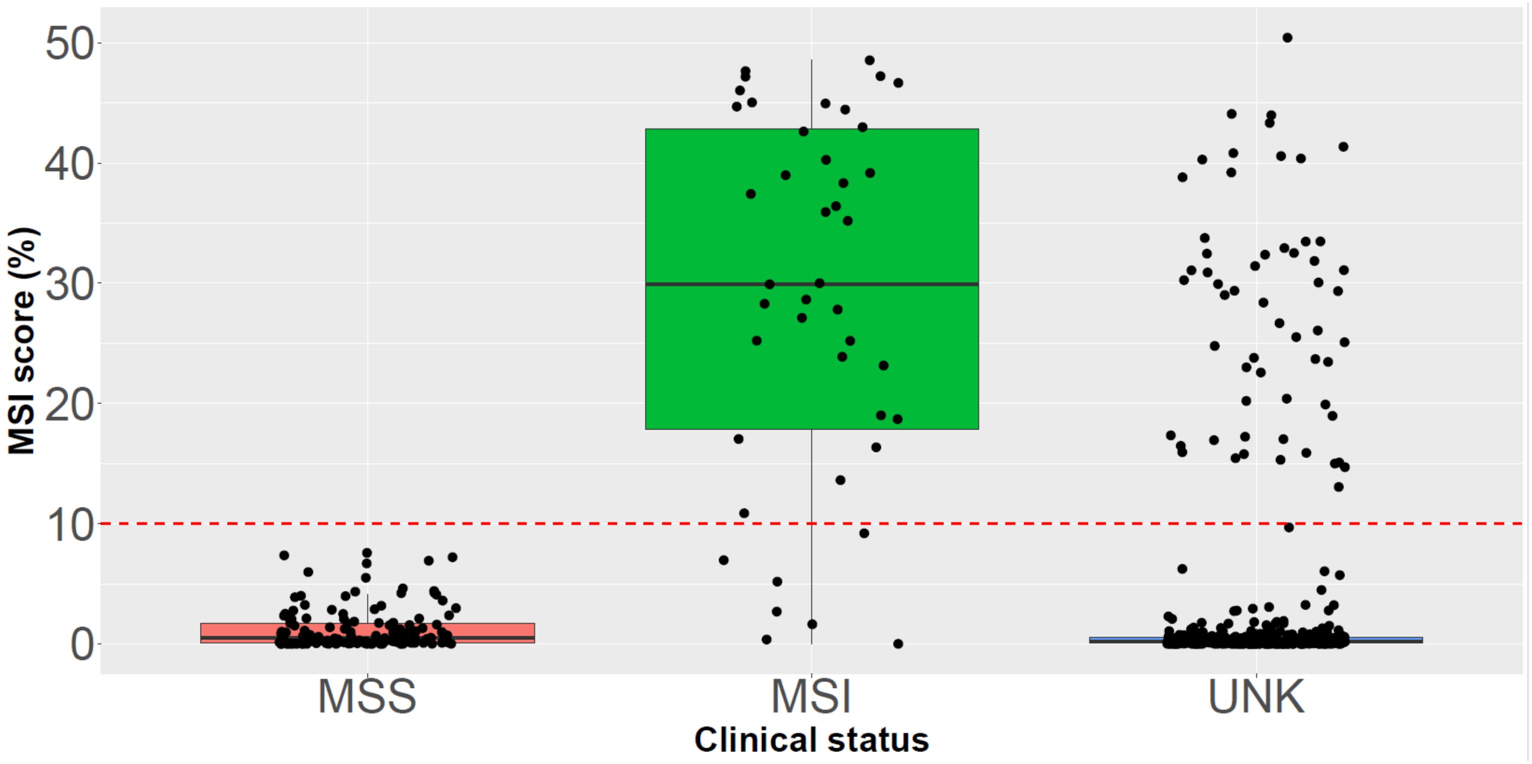


**Supplementary Figure 8.** MSI score by clinically determined dMMR/MSI status (MSS vs MSI vs unknown) in 718 colorectal cancer patients. X-axis: clinical status from medical records; Y-axis: MSI score determined by MSIsensor.
